# SARS-CoV-2 RNA is enriched by orders of magnitude in solid relative to liquid wastewater at publicly owned treatment works

**DOI:** 10.1101/2021.11.10.21266138

**Authors:** Sooyeol Kim, Lauren C. Kennedy, Marlene K. Wolfe, Craig S. Criddle, Dorothea H. Duong, Aaron Topol, Bradley J. White, Rose S. Kantor, Kara L. Nelson, Joshua A. Steele, Kylie Langlois, John F. Griffith, Amity G. Zimmer-Faust, Sandra L. McLellan, Melissa K. Schussman, Michelle Ammerman, Krista R. Wigginton, Kevin M. Bakker, Alexandria B. Boehm

## Abstract

Wastewater-based epidemiology has gained attention throughout the world for detection of SARS-CoV-2 RNA in wastewater to supplement clinical testing. Methods have been developed using both the liquid and the solid fraction of wastewater, with some studies reporting higher concentrations in solids. To investigate this relationship further, we collaborated with six other laboratories to conduct a study across five publicly owned treatment works (POTWs) where both primary solids and raw wastewater influent samples were collected and quantified for SARS-CoV-2 RNA. Solids and influent samples were processed by participating laboratories using their respective methods and retrospectively paired based on date of collection. SARS-CoV-2 RNA concentrations by mass (gene copies per gram) were higher in solids than in influent by approximately three orders of magnitude. Concentrations in matched solids and influent were positively and significantly correlated at all five POTWs. RNA concentrations in both solids and influent were correlated to COVID-19 incidence rates in the sewershed and thus representative of disease burden; the solids methods appeared to produce a comparable relationship between SARS-CoV-2 RNA concentration measurements and incidence rates across all POTWs. Solids and influent methods showed comparable sensitivity, N gene detection frequency, and calculated empirical incidence rate lower limits. Analysis of solids has the advantage of using less sample volume to achieve similar sensitivity to influent methods.

## Introduction

Wastewater represents a pooled biological sample from the contributing community and is, therefore, a resource for assessing population health. Wastewater-based epidemiology has been used to assess infectious disease burden^1–3^ and substance abuse.^4,5^ The COVID-19 pandemic has greatly increased interest in utilizing wastewater-based epidemiology to supplement clinical testing data, which can be limited due to test seeking behavior and test availability.^6^ Throughout the pandemic, researchers have successfully detected and monitored SARS-CoV-2 RNA in wastewater^7–11^ and programs have been developed to aid public health decision makers in assessing the disease burden of COVID-19 in their communities.^12,13^

Sewage consists of liquid and solid fractions, and SARS-CoV-2 RNA has been quantified in both.^8,14–16^ Sewage can be collected from publicly owned treatment works (POTWs) or from access points in the piped sewage network including at the building scale to assess disease burden.^17^ The solid fraction can be settled from raw sewage using Imhoff cones^18^ or collected from a primary clarifier, a POTW unit process that allows solids to settle as part of the treatment train.

Ye et al.^19^ previously showed that enveloped viruses partition to the solid fraction over the liquid fraction of wastewater to a greater extent than non-enveloped viruses. Motivated by this finding that solids naturally concentrate enveloped viruses, studies have compared the concentration of SARS-CoV-2 RNA in the liquid and solid components of wastewater. Li et al.^15^ compared the liquid and solid fraction of wastewater influent, using polyethylene glycol (PEG) precipitation to concentrate viruses from the liquid fraction and performing direct extraction from the solid fraction. They found that the solid to liquid SARS-CoV-2 RNA concentration ratios ranged from 10^3.6^ to 10^4.3^ mL/g. Similarly, D’Aoust et al.^14^ found higher SARS-CoV-2 RNA positivity in the solid fraction of post-grit wastewater concentrated with PEG precipitation, compared to the liquid fraction concentrated via membrane filtration. Graham et al.^8^ compared the liquid fraction of influent and solids collected from primary clarifiers at two different POTWs using a PEG concentration method for liquid influent and direct extraction for dewatered primary sludge and found the solid to liquid SARS-CoV-2 RNA concentration ratios of ∼10^3^ mL/g. Consistent with these findings, Ni et al.^16^ applied amplicon sequencing to enumerate SARS-CoV-2 genomes in sewage and noted that the solid fraction contained a considerable proportion of the viral RNA.

In this study, we compare SARS-CoV-2 RNA concentrations recovered from paired raw wastewater influent (referred to as influent in this manuscript) and primary settled solids from five different POTWs in the United States. This work is a collaborative effort among different laboratories that have retrospectively paired SARS-CoV-2 RNA data from influent and solids, some of which have been published previously.^7,20^ These data were collected as part of ongoing wastewater monitoring programs. The goal of this work is to further document differences and relationships between SARS-CoV-2-RNA measurements from the solid and liquid fraction of wastewater. We evaluate concentration ratio on a mass equivalent basis, detection frequency, and correlation with COVID-19 incident case data quantified by clinical testing. The results from this work will aid decision makers interested in utilizing SARS-CoV-2 wastewater-based epidemiology in selecting the appropriate sample matrix for their needs.

## Materials and Methods

### POTWs and method overview

Influent and settled solids samples were collected from five POTWs as part of on-going SARS-CoV-2 wastewater monitoring programs: South Bay Water Reclamation Plant (SB) in San Diego, California, USA; City of Ann Arbor Wastewater Treatment Plant (AA) in Ann Arbor, Michigan, USA; Oceanside Water Pollution Control Plant (OS) in San Francisco, California, USA; Jones Island Water Reclamation Plant (JI) in Milwaukee, Wisconsin, USA; and Orange County Sanitation District Plant 1 (OC) in Orange County, California, USA (listed in the order of size from smallest to largest). The POTWs treat average daily inflows of approximately 8, 17, 18, 75, and 120 million gallons per day (MGD) serving 125,000, 130,000, 250,000, 470,000, and 1,800,000 people in their sewersheds, respectively. All influent samples were 24-hour composites. Solid samples were taken from the primary clarifier at each POTW. Further details on sampling procedures are outlined in Table S1. Some of the POTWs add chemicals to their waste streams upstream of sample collection for odor control or improved treatment efficiency. The POTWs estimated the residence time of their primary clarifiers to be approximately between 1 to 6 hours (Table S2). Samples were collected at different intervals from April 2020 to September 2021 at cadences from daily to every other week. Influent and solids samples were matched in that they were collected on the same day. A subset of OS solids and JI influent data were previously published (Table 1).^7,20^ Here, additional OS solids and JI influent data beyond what was published previously are included.

**Table 1.** Sampling start and end dates for influent and solids at each POTW. The frequency of sampling changed over the duration of sample collection at almost every POTW, and therefore, a range is provided. Measurements obtained from a subset of the OS and JI samples have been previously published: Wolfe et al.^20^ published OS solids data from 8 Dec 2020 to 31 Mar 2021; Feng et al.^7^ published JI influent data from 30 Aug 2020 to 20 Jan 2021.

| POTW | Start Date | End Date | Frequency | Number of samples analyzed in this study |
| --- | --- | --- | --- | --- |
| SB | 4 May 2020 | 20 Nov 2020 | 3/week ~ 1/two weeks | 27 |
| AA | 22 July 2021 | 23 Sep 2021 | 3/week | 27 |
| OS | 8 Dec 2020 | 12 Jul 2021 | 1/week ~ 7/week | 101 |
| JI | 4 Aug 2020 | 26 May 2021 | 2/week ~ 1/week | 38 |
| OC | 22 Jun 2020 | 25 Nov 2020 | 2/week ~ 1/week | 23 |

Below we provide overviews of the pre-analytical processing, nucleic-acid (NA) extraction, and RNA target quantification methods used to measure SARS-CoV-2 RNA concentrations in these samples. Pre-analytical methods include all procedures used to prepare the sample for NA extraction. Analyses were carried out in six different laboratories: two processed solids samples, three influent, and one processed both. The methods varied among laboratories, but have all been described in detail in peer-reviewed publications, so brief methods are provided below with greater details in the SI. The Environmental Microbiology Minimum Information (EMMI) guidelines were followed for reporting of data.^21^

### Solids: Sample collection

Solid samples were collected by POTW staff using sterile 50-mL falcon tubes and then stored at -80°C until analysis (within 15 months - storage details shown in Table S3), with the exception of OS solids and AA solids, which were stored at 4°C and analyzed within six hours (OS) or one week (AA).

### Solids: Pre-analytical processing

Frozen solid samples were thawed at 4°C for 12-36 hours and processed according to Wolfe et al.^20^ In brief, solids were dewatered by centrifugation, then suspended in DNA/RNA shield (Zymo Research, CA) spiked with bovine coronavirus (BCoV, Calf-guard Cattle Vaccine, PBS Animal Health, OH). BCoV was used as an internal control to calculate recovery. The resuspended samples were stored at 4°C (up to 48 hours) until NA extraction. Dry weight of the dewatered solids was also determined.

### Solids: NA extraction

For AA, 0.5 g of 0.5 mm silica/zirconia beads (Biospec Products, OK) were added to each sample and homogenized by shaking with a Biospec Mini-Beadbeater-96 (Biospec Products, OK). For other POTWs, 5/32” Stainless Steel Grinding Balls (OPS Diagnostics, NJ) were added to each sample and homogenized by shaking with a Geno/Grinder 2010 (Spex SamplePrep, NJ). Nucleic acids were extracted using the Chemagic 360 and the Chemagic™ Viral DNA/RNA 300 Kit H96 (Perkin Elmer, MA). Inhibitors were removed with Zymo OneStep-96 PCR Inhibitor Removal Kits (Zymo Research, CA) before storing the RNA in -80°C for 0-78 days until analysis. Extraction negative controls (water) and positive controls (BCoV spiked in DNA/RNA shield) were included on each plate. 4 μL of Poly-A carrier RNA was added to the extraction positive controls before extraction.

### Solids: RNA target quantification

Nucleic acids were quantified through one-step droplet digital (dd)RT-PCR for SARS-CoV-2 targets (N1 and N2 at all POTWs except OS; N at OS), BCoV, and Pepper Mild Mottle Virus (PMMoV), used as a fecal strength indicator and an internal recovery control. BioRad SARS-CoV-2 droplet digital PCR kits were used with a BioRad QX200 AutoDG droplet digital PCR system (BioRad, CA). Positive and negative controls were included on all plates. Depending on the laboratory, between three and ten replicate wells were run for each sample. Results were processed using QuantaSoft and QuantaSoft Analysis Pro (BioRad, CA) to manually threshold and export data. The concentration per reaction was converted to copies per gram of dry weight using dimensional analysis (see SI). Errors are standard deviations as the “total error” from the instrument, which includes errors associated with the Poisson distribution and variability among replicate wells.

### Influent: Sample collection

Influent samples were collected by POTW staff. Large volume composite samples were collected using a 24-hour composite sampler that the POTWs already had installed onsite for routine sample collection and analysis. Aliquots of the composite samples were collected in sterile 50-ml or 500-mL bottles and stored at 4°C to be processed within 96 hours of collection (SB, AA, JI, OC); or collected in sterile 50-mL falcon tubes containing sodium chloride and buffer, stored at 4°C, and shipped or driven to the lab on ice on the day of collection to be processed within 3 days (OS).

### Influent: Pre-analytical processing

#### Filtration-based method (SB, JI, OC)

In brief, influent samples from SB and OC were acidified with 20% HCl to achieve a pH of 3.5 or lower following the methods described in Steele et al.^22^ MgCl_2_ was added to all samples to a final concentration of 25 mM, and samples were spiked with BCoV (Bovilis Coronavirus Vaccine, Merck Animal Health, NJ) as an extraction control. The samples were then filtered through 0.45-µm pore size mixed cellulose ester HA filters (Millipore Sigma, MA) or 0.8-µm pore size cellulose ester HA filters (Millipore Sigma, MA) (JI only), and the filters were stored at -80°C for between 2 hours and 2 months before NA extraction. Sterile PBS was also filtered to create a filter blank.

#### Sewage, Salt, Silica, and SARS-CoV-2 (4S) method (OS)

Samples were processed using the 4S protocol.^11^ In brief, after collection, viruses in the samples were lysed and RNA stabilized by addition of NaCl. After receiving samples in the lab, BCoV (Bovilis Coronavirus Calf Vaccine, Merck Animal Health, NJ) was spiked into the wastewater sample as a positive control and the sample was pasteurized at 70°C for 45 minutes. The sample was filtered through a 5-µm pore size PVDF filter (Millipore Sigma, MA) and the filtrate was immediately subjected to NA purification and concentration. A negative control (PBS) was also treated with the same procedure.

#### PEG precipitation method (AA)

Samples were processed according to Flood et al.^23^ using PEG to precipitate viruses. BCoV was spiked into the wastewater as a positive control and water was used as a negative control. The concentrate was used immediately for NA extraction.

### Influent: NA extraction

#### Filter-based method (SB, JI, OC)

HA filters were added to Zymo BeadBashing beads and beat for a total of two or five (JI only) minutes. After centrifuging, the supernatant was processed using a Nucleic Acid Extraction Kit (BioMerieux, NC) or an RNeasy PowerMicrobiome Kit (Qiagen, Hilden, Germany) by following the protocols provided by the manufacturers. Extraction negative controls (water or PBS) and positive controls (BCoV spiked in water or PBS) were extracted using the same protocol. Extracted NA was stored at -80°C for up to 24 hours before analysis.

#### 4S method (OS)

40 mL of 70% volume ethanol and 40 mL of filtrate were combined and processed using a Zymo III-P silica spin column (Zymo Research, CA). An extraction control (BCoV spiked in PBS) was extracted using the same protocol. The eluted RNA was stored at 4°C for same-day use or frozen at -80°C to be quantified within the next 48 hours.

#### PEG precipitation method (AA)

200 µl of sample concentrate were extracted using the QIAmp Viral RNA Mini Kit (Qiagen Sciences, MD). Extraction negative controls (water) and positive controls (BCoV) were extracted using the same protocol. RNA was used immediately for quantification.

### Influent: RNA target quantification

#### Filter-based and PEG methods (SB, AA, JI, OC)

Nucleic acids were quantified through one-step ddRT-PCR for SARS-CoV-2 (N1 and N2), BCoV, and PMMoV using the BioRad QX200 droplet digital PCR systems (BioRad, CA). Depending on the laboratory, between one and four replicates were run per sample. Positive and negative controls were included on each plate. Data was processed and exported using QuantaSoft and QuantaSoft Analysis Pro (BioRad, CA). The concentration per reaction was converted to copies per volume of wastewater using dimensional analysis. For AA, errors are standard deviations of three replicate wells. For all other POTWs, errors are the standard deviations as the “total error” from the instrument, which includes errors associated with the Poisson distribution and variability among replicate wells.

#### 4S method (OS)

SARS-CoV-2 N1, BCoV, and PMMoV were measured using one-step RT-qPCR (QuantStudio 3 Real-Time qPCR system, ThermoFisher Scientific, MA) as described by Whitney et al.^11^ Three replicates were included per sample. Negative controls were included on each plate as well as standard curves, which were also used as positive controls. Inhibition was assessed by either an internal positive control or a serial dilution, where an undiluted well was compared to a 1:5 dilution. The higher adjusted value from the comparison was used. The concentration per reaction was converted to copies per volume of wastewater using dimensional analysis.

### COVID-19 epidemiology data

For AA, laboratory-confirmed COVID-19 incident cases from residents of the city of Ann Arbor were obtained from the county health department and normalized by the city population; the sewershed is approximately defined by the city limits and it was assumed that the city level incidence rate well approximated that of the sewershed. For all other POTWs, daily counts of laboratory-confirmed COVID-19 cases with georeferenced residential address within a POTW service area shapefile were provided by the state public health department. Case data are reported as a function of the date of symptom onset (AA) or episode date (earliest of specimen collection or symptom onset date) (all other POTW). A 7-day centered moving average was calculated and used in subsequent analyses. Incidence rate was calculated using the estimated population served by each POTW.

### Statistical analysis

Statistics were computed using RStudio (version 1.4.1106). Linear regression was used to verify correlation between measurements for the two SARS-CoV-2 targets, N1 and N2. COVID-19 incidence rates were compared to SARS-CoV-2 RNA concentrations and SARS-CoV-2 RNA concentrations normalized by PMMoV concentrations.

Nonparametric Kendall’s tau and Kruskal-Wallis tests were used to assess association and significant difference between measurements, respectively, among influent and solid samples as data were neither normally nor log-normally distributed based on Shapiro-Wilk tests. To account for technical variability of wastewater measurements, Kendall’s tau was calculated using 1000 bootstrap resampling when standard deviations for the measurement were available. JI influent PMMoV, and subset of OS influent N and PMMoV were not reported with errors; therefore, raw measurement values were used without bootstrapping. Each bootstrap replicate was sampled randomly from a uniform distribution between the upper and lower bounds on the measurement. Median tau and empirical p-values were determined using the bootstrapped values.^24^ For measurements reported as non-detects (NDs), a number between zero and the lower measurement limit sampled from a uniform distribution was substituted for further analysis. Here we use the term “lower measurement limit” to represent the laboratory reported lower limit of quantification or detection (see SI). For the influent methods, each sample had a different lower measurement limit depending on the volume processed. *χ* ^2^ and Fisher Exact tests compared the frequency of non-detects.

Linear regression was used to derive slopes and y-intercepts describing empirical relationships between COVID-19 laboratory-confirmed incidence rates and measured SARS-CoV-2 gene concentrations, and between matched solids and influent measurements. Half the lower measurement limit was substituted for NDs.The lowest detectable COVID-19 incidence rate was estimated using the empirical relationships between incidence rate and SARS-CoV-2 RNA concentration at each POTW and calculating the incidence rate corresponding to the lower measurement limit reported by each participating laboratory using the *predict*.*lm* function.

## Results

### Quality assurance (QA) / quality control (QC)

Negative and positive extraction and PCR controls were negative and positive, respectively. For samples that had bovine coronavirus (BCoV) recovery quantified, recoveries suggested methods provided reasonable RNA recovery and no gross inhibition. Quantitative comparisons of BCoV recoveries were not conducted owing to complexity of interpreting surrogate recoveries.^25^

The lower measurement limits of RNA targets for solids were, on average, between ∼900 cp/g (OS) and ∼6,800 cp/g (AA); for influent they ranged, on average, from ∼0.4 cp/mL (JI) to ∼27 cp/mL (SB) (Table S4). These lower measurement limits are estimates as the exact lower measurement limit varied among samples processed since different volumes or masses were processed depending on the sample (see SI). As such, some measured concentrations could be lower than the reported average lower measurement limits.

### Measurement overview

A total of 216 pairs of matched solid and influent samples were collected from five POTWs. Across solids samples, PMMoV ranged from 9.7 × 10^7^ to 6.8 × 10^9^ copies/g of dry weight (median = 5.9 × 10^8^); across influent samples, PMMoV ranged from 6.7 × 10^2^ to 2.7 × 10^6^ copies/mL of wastewater (median = 6.9 × 10^4^) (Figure S1). PMMoV was different between POTWs (Kruskal-Wallis P < 10^−15^) within the same matrix (i.e., solid or influent); OS tended to have lower PMMoV than other POTWs in solids (by 0.2 - 0.8 log units), and OS and JI had lower PMMoV than other POTWs in influent (by 1 - 1.5 log units). The median ratio of PMMoV concentrations in matched solids to influent samples across all POTWs was 6×10^3^ (n = 207, 9 influent samples with no PMMoV measurements were omitted; range 4×10^2^ to 3×10^5^). Ratios were statistically different between POTWs with JI having the highest median ratio (median = 3×10^4^) and SB and OC the lowest (median = 1×10^3^) (Kruskal-Wallis P < 10^−15^) (Table S5).

In solids, N1, N2, and N gene targets were measured; the N target is located in approximately the same location in the SARS-CoV-2 genome as the N1 target.^26^ SARS-CoV-2 RNA gene concentrations in solids ranged from ND to 2.4 × 10^6^ cp/g dry weight (N1 or N) and ND to 2.1 × 10^6^ cp/g dry weight (N2). Across influent samples, SARS-CoV-2 RNA concentrations ranged from ND to 7.3 × 10^2^ cp/mL (N1) and from ND to 1.2 × 10^3^ cp/mL (N2) (Figure S2). Across all solids measurements, N1 and N2 were strongly and positively correlated (R^2^ = 0.99, slope = 1.1, p-value < 10^−15^) (Figure S3). Similarly, across all influent measurements, N1 and N2 were strongly and positively correlated (R^2^ = 0.94, slope = 0.6, p-value < 10^−15^) (Figure S4). Therefore, further analyses focused on the N assay for OS solids and the N1 assay for all other samples (Figure 1). All wastewater data presented in the paper is publicly available through the Stanford Digital Repository (https://purl.stanford.edu/kd763fh7892).

**Figure 1.**
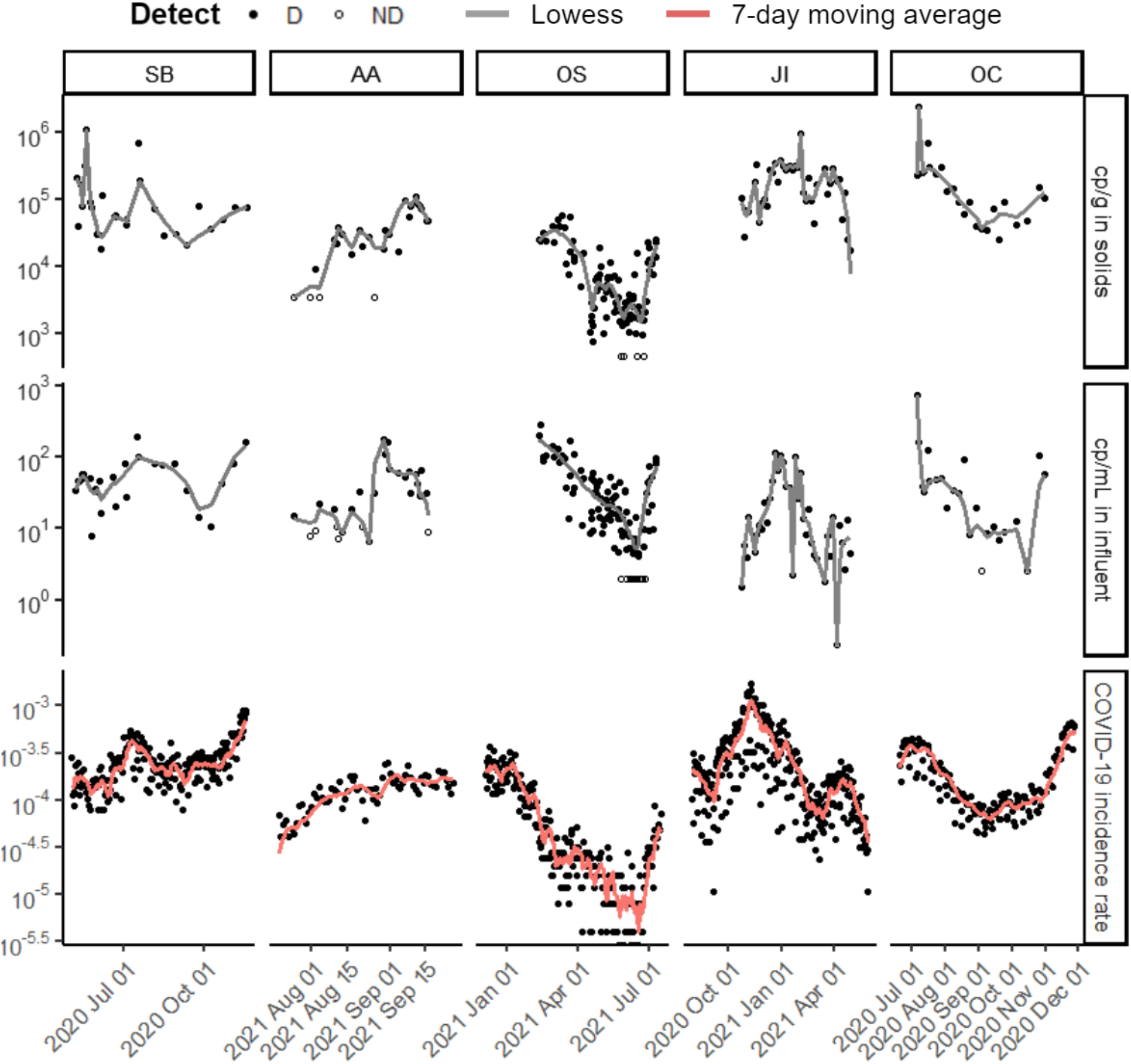
Time series of (top to bottom) SARS-CoV-2 targets N1 or N measured in solids (cp/g dry weight), concentration measured in influent (cp/mL), and laboratory-confirmed SARS-CoV-2 incidence rate for each of the five POTWs over their respective duration of sample collection. N was measured for OS solids and N1 for all other data sets. Each wastewater data point represents SARS-CoV-2 RNA concentration for a single sample as reported by the respective laboratory. Replication was performed differently for each lab (see SI). Samples above the lower measurement limit are shown as filled circles. Samples that resulted in ND, shown as empty circles, were substituted with a value half of the sample’s lower measurement limit. Lines for solids and influent are locally weighted scatterplot smoothing (lowess) with value of **α** that minimizes the residual for each dataset (Table S6)^33^. Lines for clinical are 7-day centered smoothed averages. The same time series with normalization by PMMoV can be found in the SI (Figure S5).

### Relationship between SARS-CoV-2 RNA in solids and influent

The median ratio of SARS-CoV-2 RNA concentrations in matched solids and influent across all POTWs was 9×10^2^ (n = 216, 25th percentile 3×10^2^, 75th percentile 4×10^3^). Ratios were statistically different between POTWs, with JI having the highest median ratio and OS the lowest (Kruskal-Wallis P < 10^−15^, Table 2). SARS-CoV2 RNA concentrations in matched solids and influent were positively and significantly correlated at all five POTWs as both aggregated data (Figure 2, median Kendall’s tau = 0.22, empirical p-value < 0.001) and at individual plant level (Table S7). To derive an empirical relationship between the log_10_-transformed solids and liquid concentrations, we used linear regression where Y is the log_10_-transformed solids concentration (cp/g) and X is the log_10_-transformed influent concentration (cp/mL) consistent with a Freundlich isotherm model, assuming influent concentrations are representative of concentrations in the liquid fraction.^27^ Slopes ranged from 0.26 to 0.63 and y-intercepts ranged from 2.89 to 4.89 (Table S8) depending on the POTW, consistent with n = 2 to 3, and K_f_ = 10^3^ - 10^5^ ml/g in the Freundlich model: *C*_*s*_ = *K*_*f*_ *C*_*l*_^*1/n*^ where *C*_*s*_ is RNA concentration in solid fraction, *C*_*l*_ is RNA concentration in liquid fraction, *K*_*f*_ is Freundlich’s constant, and 1/*n* is the exponent of non-linearity.

**Table 2.** SARS-CoV-2 RNA gene concentration ratios in matched solids to influent for the five POTWs, listed as rows. The ratios were calculated on an equivalent mass basis for N1 or N. For samples that resulted in ND, half of the lower measurement limit was used. Number of matched samples and 25th percentile, median, and 75th percentile ratios calculated for the plants are reported.

| **POTW** | **n** | **25th percentile**<br>**(mL/g)** | **Median**<br>**(mL/g)** | **75th percentile**<br>**(mL/g)** |
| --- | --- | --- | --- | --- |
| SB | 27 | 860 | 1,400 | 3,600 |
| AA | 27 | 380 | 1,100 | 2,000 |
| OS | 101 | 130 | 280 | 530 |
| JI | 38 | 4,700 | 10,000 | 20,000 |
| OC | 23 | 3,100 | 5,500 | 7,500 |

**Figure 2.**
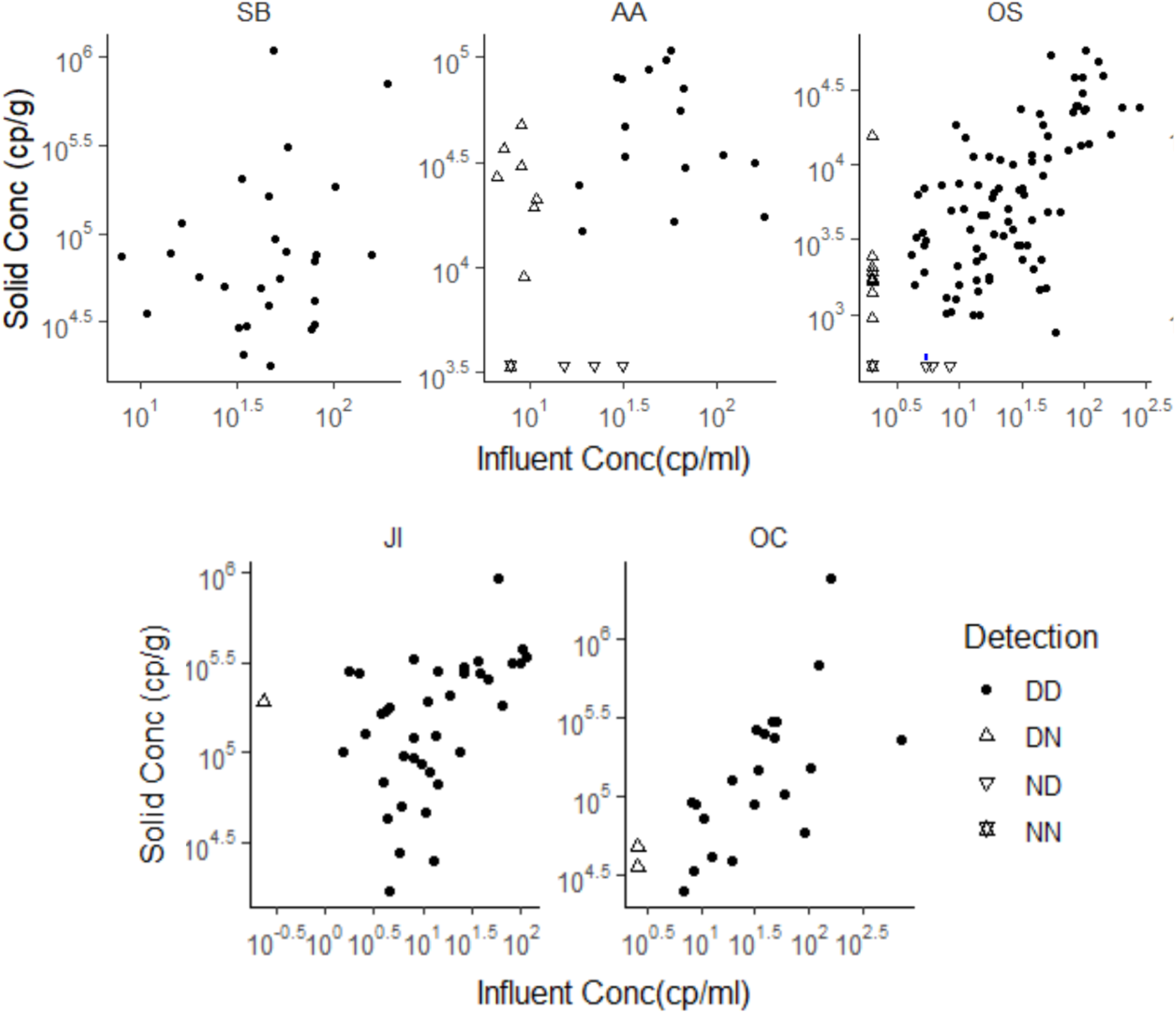
SARS-CoV-2 RNA concentrations in matched solid and influent samples. N1 concentration was used for this analysis, with the exception of OS solids where concentration of N was used. Each data point represents SARS-CoV-2 RNA concentration for a single sample as reported by the respective laboratory. Replication was performed differently for each lab (see SI). All data above its lower measurement limit are shown as filled circles. Data points with ND in influent are shown as an empty upright triangle, points with ND in solids are shown as an empty upside-down triangle, and points where both were ND are shown as empty overlapped upright and upside-down triangles. NDs have been substituted as half of the sample’s lower measurement limit. Note that the data are displayed in log_10_-scale format for ease of visualization.

Detection frequency was calculated for matched solids and influent samples, along with empirical incidence rate lower limit for all samples (Table 3). Overall detection frequency of N1 or N was 96% for solids and 90% for influent: there were eight of 216 solids samples and twenty-one of 216 influent samples that resulted in ND for N1 or N. The frequency of NDs in solids and influent were not significantly different (chi-square test or fisher exact test, p > 0.05). Detection limit in terms of incidence rate was similar between solids and influent at all POTWs: in solids, the limit ranged from 0.7 to 20 out of 100,000, and in influent, the limit ranged from 0.9 to 18 out of 100,000. Over the duration of the study, the lowest 7-day smoothed incident rates observed in each plant ranged from 0.4 to 12 cases per 100,000 at OS and SB, respectively.

**Table 3.** Detection frequency (“Frequency”) and incidence rate limit (“Limit”) for samples from the five POTWs. Detection frequency denotes how many samples were above the lower measurement limit. Incidence rate limit is the incidence rate (out of 100,000) corresponding to the average SARS-CoV-2 RNA lower measurement limit as modeled using linear regression. Errors on the detection limit represent the standard error on the prediction.

|  | SB |  | AA |  | OS |  | JI |  | OC |  |
| --- | --- | --- | --- | --- | --- | --- | --- | --- | --- | --- |
|  | Solid | Influent | Solid | Influent | Solid | Influent | Solid | Influent | Solid | Influent |
| **Frequency** | 27/27 | 27/27 | 23/27 | 19/27 | 97/101 | 91/101 | 38/38 | 37/38 | 23/23 | 21/23 |
| **Limit**<br>**(#/100,000)** | 20±4 | 18±1 | 11±1 | 13±1 | 0.7±0.1 | 0.9±0.1 | 5±2 | 5±1 | 3±1 | 8±2 |

### Relationship between SARS-CoV-2 RNA in wastewater solids and incidence rates

SARS-CoV-2 RNA concentrations in solids correlated positively and significantly to COVID-19 incidence rates. Kendall’s tau between 7-day smoothed incidence rates and SARS-CoV-2 RNA concentrations in solids ranged from 0.07 (SB) to 0.56 (OC) (median = 0.36, empirical p-value < 0.005 for all) (Table 4). Linear regression was used to derive an empirical relationship between log_10_-transformed COVID-19 incidence rate and log_10_-transformed solid concentration. The regression showed that for 1 log10 increase in SARS-CoV-2 N1 or N cp/g, there was between 0.02 and 0.75 log_10_ increase in incidence rate; there was a similar positive log_10_ increase when data were normalized by PMMoV (Figure S5, Table 5). The data from all five POTWs appear to fall on a single line (Figure 3) when plotted as COVID-19 incidence rate versus SARS-CoV-2 RNA concentration (median tau = 0.64, p < 0.001) or SARS-CoV-2 RNA concentration normalized by PMMoV (median tau = 0.58, p < 0.001); when data are concatenated and analyzed together, the slope of the regression suggests that a 1 log_10_ increase in SARS-CoV-2 N1 or N cp/g corresponds to a 0.62 (± 0.02 standard error) log_10_ increase in incidence rate (R^2^ = 0.70, p-value < 10^−15^); for concentration normalized by PMMoV, there is a 0.64 (± 0.03 standard error) log_10_ increase in COVID-19 incidence (R^2^ = 0.61, p-value < 10^−15^).

**Table 4.** Median Kendall’s tau correlation between wastewater SARS-CoV-2 RNA N gene concentration (N1 or N) and incidence rate in each sewershed. 1000 instances of Kendall’s tau were calculated by bootstrapping upper and lower confidence intervals for measured concentration of SARS-CoV-2 RNA. Confidence intervals were not available for all OS influent samples, and therefore Kendall’s tau was calculated with raw data points. For samples that resulted in ND, the lower measurement limit and 0 were used as upper and lower confidence intervals respectively. Kendall’s tau was calculated with raw N gene wastewater concentration and with values normalized by PMMoV. Empirical p-value was lower than 0.005 for all unless otherwise stated in parenthesis.

| Plant | Solid |  | Influent |  |
| --- | --- | --- | --- | --- |
|  | N1 or N | N1/PMMoV or N/PMMoV | N1 or N | N1/PMMoV or N/PMMoV |
| All | 0.64 | 0.58 | 0.24 | -0.03<br>(p-value = 1) |
| SB | 0.07 | 0.11 | 0.33 | 0.21 |
| AA | 0.37 | 0.46 | 0.40 | 0.34 |
| OS | 0.52 | 0.61 | 0.60 | 0.47 |
| JI | 0.36 | 0.20 | 0.52 | 0.48 |
| OC | 0.56 | 0.54 | 0.51 | 0.68 |

**Table 5.**
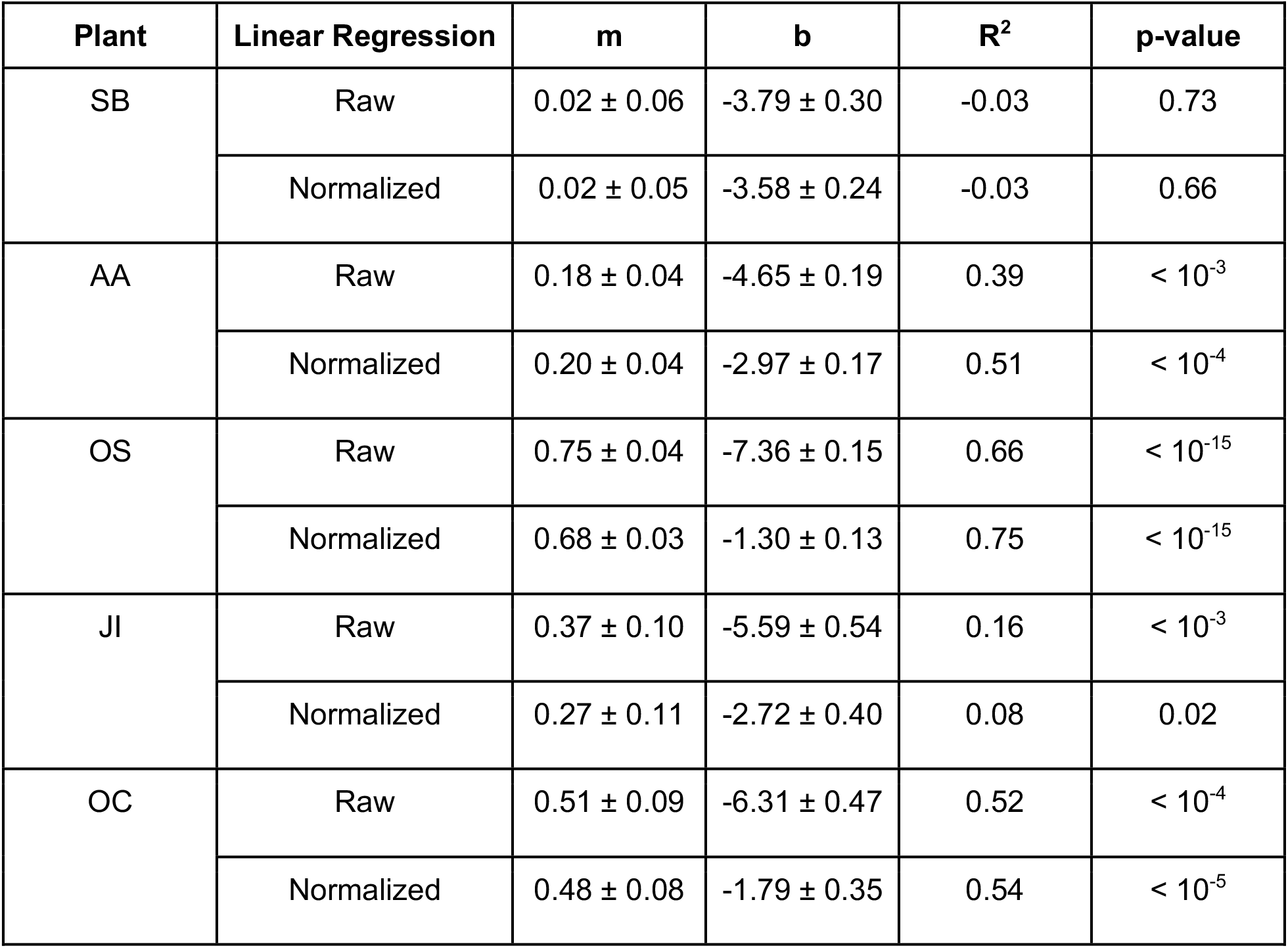
Empirical relationship between SARS-CoV-2 RNA N gene (N1 or N) concentrations measured in solids and COVID-19 incidence rates. Coefficients are presented for linear regression to raw data and data normalized by PMMoV. Y = mx + b where y = log_10_-transformed COVID-19 incidence rates, m = slope, b = intercept, and x = log_10_-transformed solids concentration. The error on m and b represents standard error for the calculated coefficients. R^2^ and p-value are provided for completeness but the regression is used to derive an empirical relationship between the variables; to assess association, Kendall’s tau was used (see Table 4).

| Plant | Linear Regression | $m$ | $b$ | $R^2$ | $p$-value |
| --- | --- | --- | --- | --- | --- |
| SB | Raw | $0.02 \pm 0.06$ | $-3.79 \pm 0.30$ | -0.03 | 0.73 |
| | Normalized | $0.02 \pm 0.05$ | $-3.58 \pm 0.24$ | -0.03 | 0.66 |
| AA | Raw | $0.18 \pm 0.04$ | $-4.65 \pm 0.19$ | 0.39 | $< 10^{-3}$ |
| | Normalized | $0.20 \pm 0.04$ | $-2.97 \pm 0.17$ | 0.51 | $< 10^{-4}$ |
| OS | Raw | $0.75 \pm 0.04$ | $-7.36 \pm 0.15$ | 0.66 | $< 10^{-15}$ |
| | Normalized | $0.68 \pm 0.03$ | $-1.30 \pm 0.13$ | 0.75 | $< 10^{-15}$ |
| JI | Raw | $0.37 \pm 0.10$ | $-5.59 \pm 0.54$ | 0.16 | $< 10^{-3}$ |
| | Normalized | $0.27 \pm 0.11$ | $-2.72 \pm 0.40$ | 0.08 | 0.02 |
| OC | Raw | $0.51 \pm 0.09$ | $-6.31 \pm 0.47$ | 0.52 | $< 10^{-4}$ |
| | Normalized | $0.48 \pm 0.08$ | $-1.79 \pm 0.35$ | 0.54 | $< 10^{-5}$ |

**Figure 3.**
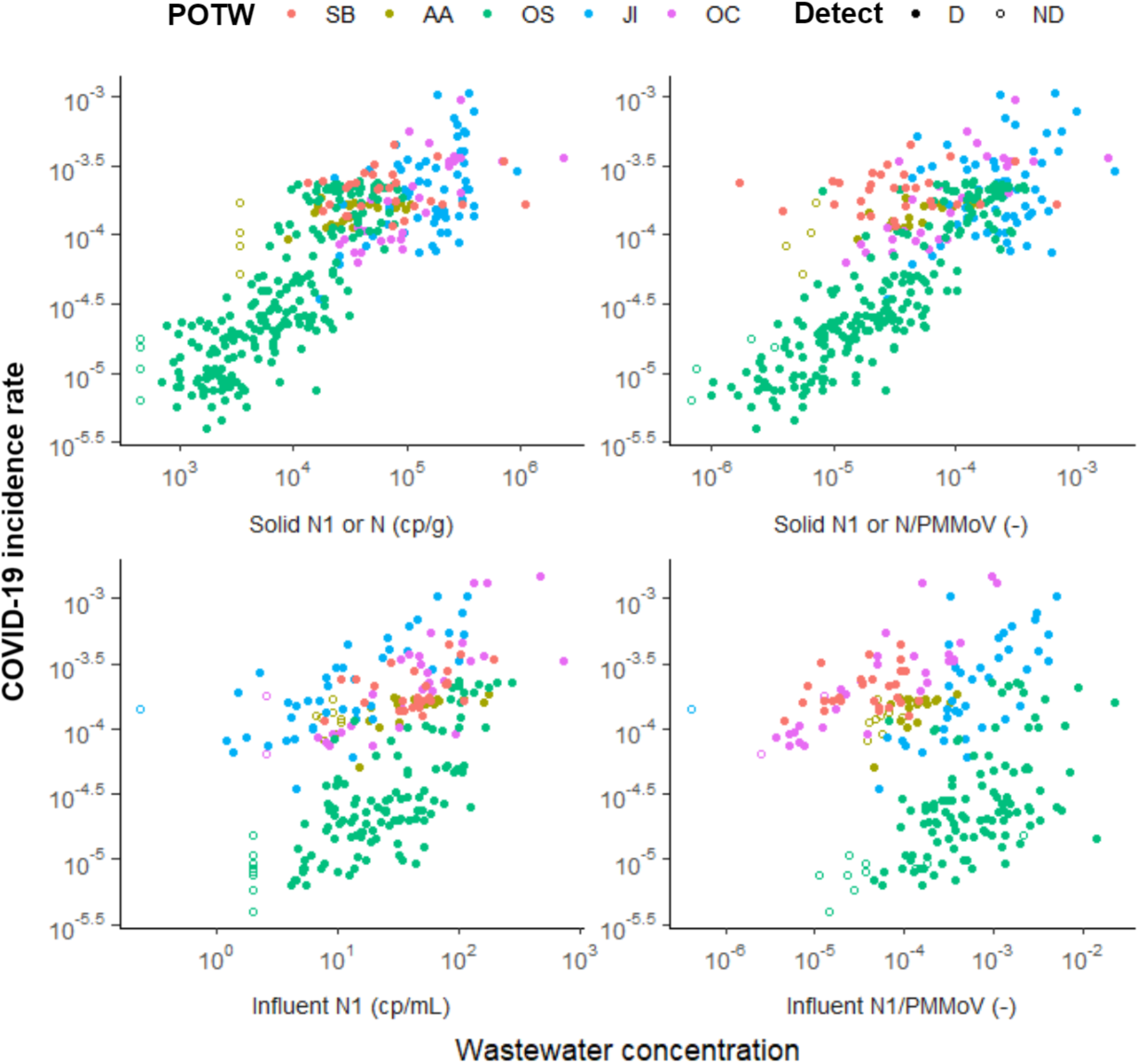
7-day smoothed COVID-19 incidence rate plotted against SARS-CoV-2 concentration in solids (top row) and influent (bottom row). From left to right, plots show the association between incidence rate and N1 or N; and N1 or N normalized by PMMoV for samples that had corresponding values of PMMoV. Samples above the lower measurement limit are shown as filled circles. Samples that resulted in ND, shown as empty circles, were substituted with a value half of the sample’s lower measurement limit.

### Relationship between SARS-CoV-2 RNA in wastewater influent and incidence rates

SARS-CoV-2 RNA measurements from influent positively and significantly correlated to COVID-19 incidence rates. Kendall’s tau between 7-day smoothed incidence rates and SARS-CoV-2 RNA concentrations in influent ranged from 0.33 (SB) to 0.60 (OS) (median = 0.51, empirical p-value < 0.005) (Table 4). Linear regression between log_10_-transformed COVID-19 incidence rate and log_10_-transformed influent concentration showed that for 1 log_10_ increase in N1 concentration, there is between a 0.18 and 0.62 log_10_ increase in incidence rate across different POTWs. There was a similar positive log_10_ increase when data was normalized by PMMoV (Figure S6, Table 6). When data from the five POTWs are concatenated and analyzed together, there is a positive association between incidence rate and SARS-CoV-2 RNA concentration (tau = 0.24, p < 0.001); but not for concentration normalized by PMMoV (tau = -0.03, p = 1) (Figure 3). Linear regressions suggest a 1 log increase in N was associated with a 0.49 ± 0.06 log increase in incidence rate (R^2^ = 0.22, p < 10^−14^); for N normalized by PMMoV a 1 log increase in N1/PMMoV corresponds to a 0.03 reduction in incidence rate (slope = -0.03 ± 0.05, R^2^ = -0.003, p = 0.58).

**Table 6.** Relationship between SARS-CoV-2 RNA N gene (N1 or N) measured in influent and COVID-19 incidence rates. Y = mx + b where y = log_10_-transformed COVID-19 incidence rates, m = slope, b = intercept, and x = log_10_-transformed influent concentration. The error on m and b represents standard error. R^2^ and p-value are provided for completeness, but the regression is used to derive an empirical relationship between the variables; to assess association, Kendall’s tau was used (see Table 4).

| Plant | Linear Regression | $m$ | $b$ | $R^2$ | $p$-value |
| --- | --- | --- | --- | --- | --- |
| SB | Raw | $0.18 \pm 0.09$ | $-4.00 \pm 0.14$ | 0.12 | 0.04 |
| | Normalized | $0.09 \pm 0.07$ | $-3.29 \pm 0.30$ | 0.03 | 0.18 |
| AA | Raw | $0.18 \pm 0.05$ | $-4.12 \pm 0.07$ | 0.31 | $< 10^{-2}$ |
| | Normalized | $0.28 \pm 0.07$ | $-2.74 \pm 0.28$ | 0.36 | $< 10^{-3}$ |
| OS | Raw | $0.62 \pm 0.05$ | $-5.40 \pm 0.07$ | 0.57 | $< 10^{-15}$ |
| | Normalized | $0.37 \pm 0.05$ | $-3.39 \pm 0.17$ | 0.32 | $< 10^{-10}$ |
| JI | Raw | $0.42 \pm 0.06$ | $-4.12 \pm 0.08$ | 0.50 | $< 10^{-7}$ |
| | Normalized | $0.29 \pm 0.07$ | $-2.74 \pm 0.24$ | 0.29 | $< 10^{-3}$ |
| OC | Raw | $0.48 \pm 0.08$ | $-4.42 \pm 0.14$ | 0.53 | $< 10^{-5}$ |
| | Normalized | $0.42 \pm 0.05$ | $-1.81 \pm 0.21$ | 0.74 | $< 10^{-9}$ |

## Discussion

We compared measurements of SARS-CoV-2 RNA in solids and raw wastewater influent from five POTWs. Across all matched solids and influent samples, the median ratio of SARS-CoV-2 RNA in solids to influent was ∼10^3^. This result suggests that SARS-CoV-2 RNA, present in virions, fragmented virions, or outside of virions,^28^ partitions to the solid fraction of wastewater. We also found that PMMoV RNA is enriched in the solids fraction relative to influent; the median ratio of PMMoV RNA in solids to influent was ∼10^4^. These results support earlier findings that SARS-CoV-2 RNA is enriched in the solids fraction of wastewater by 3-4 orders of magnitude on a mass equivalent basis.^8,14,15^ Previous reports suggest that other viruses and bacteriophages also have a high affinity for wastewater solids including enteroviruses, rotavirus, murine hepatitis virus, phi 6, and adenovirus.^19,29–31^ However, given the heterogeneity of virus capsid structures, more research is needed to identify whether there are viruses that do not partition to solids.

The settled solids collected in this study for analysis entered the POTWs as solids suspended in the influent, and then settled as primary sludge in the primary clarifier. The suspended solids content of influent is typically on the order of 10^2^ mg/L. Assuming that the concentration of SARS-CoV-2 RNA in settled solids is representative of its concentration in suspended solids, and that solids contain three orders of magnitude more SARS-CoV-2 RNA than influent, the concentration of suspended solids in influent contributes only 10% to the total amount of SARS-CoV-2 RNA in influent. Therefore, the majority of SARS-CoV-2 RNA measured in influent is from the liquid phase even when suspended solids are retained in the measurement method.

The ratio of concentrations in solids and influent can be conceptualized as an empirical partitioning coefficient K_d_, assuming the majority of SARS-CoV-2 measured in influent is present in the liquid phase. K_d_ varied among samples and POTWs. Partitioning characteristics may be influenced by properties of the solid and liquid matrix in the mixture. For example, partitioning of organic chemicals is controlled in part by the organic carbon and mineral content of the solid matrix, the ionic strength of the liquid, pH, and temperature.^27^ Given the complex and variable nature of wastewater, it is not surprising that K_d_ varies in matched samples among and between POTWs. To investigate how K_d_ varies as a function of the solids characteristics, we compared K_d_ to PMMoV in solids. Here we used PMMoV as a proxy for the fecal strength of the solids, and therefore as a measure of organic content of the solids. We found that K_d_ is positively and significantly associated with solids PMMoV concentration (Kendall’s tau = 0.4, p<10^−14^). Additional work will be needed to better understand what controls partitioning of viruses to solids in wastewater and whether a partitioning model, which requires an equilibrium assumption, is appropriate.

We also must consider the possibility that K_d_ is affected by the approaches used to obtain and measure SARS-CoV-2 RNA in the solid and liquid matrices. All of the solids approaches were similar in their pre-analytical and NA extraction approaches because RNA is already concentrated in a small volume of sample: dewatered solids were suspended in a solution, and NA were extracted directly from a small volume (<1 ml) of this solution using commercial NA extraction kits. In contrast, the influent approaches had diverse pre-analytical and NA extraction steps that involved collecting and concentrating SARS-CoV-2 RNA from a large (>20 ml) volume of liquid. K_d_ might be lower when influent SARS-CoV-2 is measured with an approach that is more efficient at recovering SARS-CoV-2 RNA from influent than others. Interestingly, the lowest K_d_ values were observed at OS, the only plant that used the 4S method. To determine how influent methods compare and whether the low K_d_ values observed at OS can be attributed to the method used to measure SARS-CoV-2 RNA in the influent, additional measurements of SARS-CoV-2 RNA in matched solids and influent using the 4S and other influent methods would need to be collected. It is also necessary to acknowledge that although we did our best to match solids and influent samples while taking advantage of ongoing wastewater-based epidemiology sampling efforts, the matching approach is imperfect. For example, solids samples are akin to a 1 to 24 hour composite samples, depending on the collection approach, based on estimations of solids residence time of primary clarifiers provided by POTW staff. On the other hand the influent samples were 24-hour composite samples. In the future, researchers could investigate partitioning of SARS-CoV-2 RNA or other viruses by collecting a large volume of influent that is then split into (1) a sample to be processed using an influent method, and (2) a sample to be settled in an Imhoff cone^18^ then processed using a solids method. It is important to acknowledge that samples were archived and stored in different ways for different durations; this may also have impacted the enumeration of SARS-CoV-2 RNA; however, each lab followed best practices using storage methods that they have tested previously.^11,22,32^

In order to compare the sensitivity of the solids and influent measurements, we determined the COVID-19 incidence rate below which we expect the measurements to yield non-detects. This was accomplished by deriving an empirical relationship between SARS-CoV-2 RNA concentrations and incidence rates at each POTW for solids and influent measurements then calculating the incidence rate corresponding with the average lower measurement limit for the method. Solids and influent methods yielded similar sensitivity across POTWs. Both were able to detect SARS-CoV-2 RNA when incidence rates were between ∼1 and ∼10/100,000. Influent and solids measurements were the most sensitive at OS where they could detect <1/100,000 incidence rate. It is not clear at the present time what sensitivity is needed for wastewater monitoring to be informative for pandemic response. Given that the size of the sewersheds range from 10^5^ to 10^6^ people, it appears wastewater monitoring using these methods can reliably identify when there are between 1 and 100 people in the sewershed with laboratory-confirmed COVID-19, depending on the POTW. The lower measurement limits of these methods may be reduced further, should public health officials determine that a lower incidence rate threshold is needed to guide public health recommendations.

In a previous study, we suggested that methods for detecting SARS-CoV-2 RNA in wastewater at POTWs should be *representative, comparable, sensitive*, and *scalable* in order to provide actionable insight on COVID-19 incidence.^20^ *Representative* means that the measurements correlate with COVID-19 incidence. In this study, measurements in solids and influent both are positively associated with COVID-19 incidence, and the positive association held when SARS-CoV-2 measurements were normalized by PMMoV. The magnitude of association varied across POTWs similar to results reported by others.^7,20,24,33^ The weakest association was observed at SB for both solids and influent. The reasons why associations were weakest at this POTW are unknown, but could be due to the relatively static COVID-19 incidence, which changed by less than one order of magnitude over the duration of sampling or because COVID-19 case data were unreliable early in the pandemic when many of the SB samples were collected. It is also important to note that COVID-19 case data likely under-represent the actual number of infections in the sewersheds^34^ and this may vary among locations and across times, which would affect the associations between incidence rate and wastewater concentrations of SARS-CoV-2 RNA. The apparent power-law relationship between SARS-CoV-2 RNA concentrations and incidence rates is consistent with under-reporting of COVID-19 cases when incidence rates are high.^35^

*Comparable* means that samples measured at different POTWs and by different labs can be combined and compared to infer relative incidence rates across communities within POTW service areas. Solids data from the five POTWs from different regions of the United States appear to collapse on a single curve when plotted as incidence rate versus SARS-CoV-2 RNA concentration suggesting that a 1 log_10_ increase on SARS-CoV-2 concentrations corresponded to a 0.6 log_10_ increase in incidence rates; this relationship is similar to those published by Wolfe et al.^20,24^ using different solids data sets that were obtained using different approaches and laboratories. This previous work showed how measurements of SARS-CoV-2 RNA in solids obtained using different pre-analytical methods could be scaled by PMMoV to be comparable.^20^ Influent data from the five POTWs do not visually appear to fall on the same curve when plotted as incidence rate versus SARS-CoV-2 RNA concentration, perhaps because the different influent methods are not themselves comparable. Different influent methods likely recover different fractions of the SARS-CoV-2 signal;^33^ at the same incidence rate, a higher wastewater concentration was reported for OS, the only POTW monitored with the 4S method. When influent data from different methods were scaled by PMMoV, the data again did not appear to fall on a single curve. Based on the results of the present study and previous work, solids measurements appear to be comparable. However, the influent measurements presented herein were not comparable. Additional work is needed to better understand how to scale influent measurements obtained from different POTWs or how to normalize diverse influent methods so that they can be compared and used to infer relative incidence rates across sewersheds.

*Sensitive* describes the lower detection limits of the methods. Numerically, the lowest measurable concentration for the solids methods are higher than the influent methods, but it is inappropriate to compare these numbers directly because they have different units and the target undergoes partitioning to the solid phase. For the solids methods, the smallest lower measurement limits were obtained from methods that merged the largest number of wells during digital PCR: OS merged ten wells, JI, SB, OC merged six wells, and AA used three wells. Decreasing the lower measurement limit within the solids methods is possible and can be accomplished by increasing the mass of solids suspended per mL in the DNA/RNA shield solution prior to extraction or increasing the number of wells merged. The challenge with the former is that increasing solids concentrations can increase inhibition of the RT-PCR while the challenge with the latter is increasing reagent costs. Within the influent methods, the smallest lower measurement limit was achieved using the JI membrane filtration method and the largest lower measurement limit was achieved using the SB membrane filtration method (both with digital PCR) due to different effective volume processed. Decreasing the lower measurement limits of the influent methods is possible and would require increasing the volume of influent processed in the pre-analytical methods, or increasing the number of merged wells during digital PCR. OS influent samples were the only samples processed by qPCR, and the lower measurement limit could potentially be decreased by using digital PCR. Increasing the influent volume processed may increase inhibition of the RT-PCR and can be difficult or impossible using dead-end filtration due to filter clogging. Wastewater is a complex and variable matrix with a wide range of RT and PCR inhibitory substances including organic and inorganic molecules.^36,37^ Future work to characterize and alleviate RT and PCR inhibition using different NA extraction kits, inhibitor removal kits, or mastermixes as well as testing methods that concentrate fewer PCR inhibitory substances, is warranted to improve sensitivity of both solids and influent methods.

*Scalable* means that methods are amenable to automation and high-throughput processing with the use of automated instruments and liquid handling robots that generate results quickly (i.e., on the day of receiving a sample). The solids methods implemented in this study are scalable; the OS data were generated using automated NA extraction systems, liquid handling robots, and digital PCR methods with results available the same day as sample collection. Measurement of percent solids of each sample may be hard to automate but can be skipped if the final reported RNA concentration is normalized by PMMoV concentration. The influent methods used in this study were not executed in an automated, high throughput format and may be difficult to scale. All require volumes greater than 10 mL and include time and staff-intensive filtration or flocculation steps. The limited influent methods that are scalable use small volumes (at most 10 mL),^38^ which limits the sensitivity of the methods.

## Conclusion

SARS-CoV-2 RNA in wastewater measured using the diverse methods described in this study are representative of COVID-19 incidence and were adequately sensitive to detect the virus when incidence rates were low (∼1/10^5^). SARS-CoV-2 RNA and PMMoV RNA were enriched in solids relative to influent (on a per mass basis) and thus solids naturally concentrated the viral targets. Owing to the lower concentrations of SARS-CoV-2 RNA in influent, large influent volumes must be processed prior to analysis unless community disease burdens are very high. Multiple effective methods for recovering viruses from liquid wastewater are described in this paper, but each likely recovers different fractions of the SARS-CoV-2 RNA signal at different efficiencies and thus are difficult to compare to one another. Since SARS-CoV-2 RNA naturally concentrates in solids direct extraction of nucleic acids from small masses of solids is possible and an effective way to measure SARS-CoV-2 even when disease burdens are low (<1/10^5^). All methods were representative and sensitive, and methods based on solids appear to also be comparable across POTWs and variations in pre-analytical methods, and scalable to a high throughput, robotic format. Further work should be done to determine if these advantages can be realized in SARS-CoV-2 RNA measured in influent methods. Both solids and influent methods can be made more sensitive by altering methods, but inhibition during quantification may represent an obstacle to doing so.

## Data Availability

All wastewater data are available in the Stanford Digital Repository and are free to download to the public.

https://purl.stanford.edu/kd763fh7892

## Acknowledgements

This work was partially supported by the CDC Foundation and an NSF RAPID (CBET-2023057) to K.R.W. and A.B.B, in addition to Stanford Campus Incheon Global Campus and Stanford School of Engineering Graduate Fellowship to S.K. K.M.B. was partially supported by NIH awards KL2TR002241 & UL1TR002240. We also thank the Milwaukee Metropolitan Sewerage District for a Graduate Student Fellowship for M.K.S. and the Catena Foundation for a grant to K.L.N. and R.S.K. We thank the California Department of Public Health COVID-19 Wastewater Surveillance, Epidemiology and Data teams; Wisconsin DHS (Jonathan Meiman and Nathan Kloczko); and Washtenaw County Health Department for the COVID-19 incidence data. Numerous people contributed to the sample collection. We thank the participating POTWs for providing the samples and their expertise on individual systems, including Lily Chan, Alexander Miot, Ryan Batjiaka, and the Oceanside plant operations personnel (OS); Margil Jimenez and others from OCSan team (OC); Ernesto Molas, Steven Bates, Rowena Ronsairo, and South Bay Wastewater Chemistry Laboratory (SB); Sharon Mertens and Matt Magruder (JI); and Kyle Weinman (AA). We are grateful to Matt Metzger, Melissa Thornton, and Justin Paluba at UC Berkeley; David Catoe, Archana Anand, and Winnie Zambrana at Stanford; Madison Griffith, Lucy Mao, Jeffrey Chokry, Eric Macias, and David Wanless at Southern California Coastal Water Research Project for their work in processing samples.

## Author Contributions

**Sooyeol Kim:** Conceptualization, Methodology, Validation, Investigation, Formal analysis, Data curation, Writing - original draft, Visualization. **Lauren C. Kennedy:** Methodology, Formal analysis, Data curation, Writing - original draft, Visualization. **Marlene K. Wolfe:** Data curation, Conceptualization, Writing - review & editing. **Craig Criddle:** Writing - review & editing, Supervision. **Dorothea H. Duong:** Data Curation, Methodology, Validation, Writing - review & editing. **Aaron Topol:** Data curation, Methodology, Validation, Writing - review & editing. **Bradley J. White:** Resources, Data Curation, Methodology, Validation, Writing - review & editing. **Rose S. Kantor:** Data curation, Formal analysis, Writing - review & editing. **Kara L. Nelson:** Writing - review & editing, Supervision, Resources, Funding acquisition. **Joshua A. Steele:** Methodology, Validation, Writing - review & editing. **Kylie Langlois:** Data Curation, Methodology, Validation, Writing - review & editing. **John F. Griffith:** Resources, Writing - review & editing, Supervision. **Amity G. Zimmer-Faust:** Data Curation, Methodology, Validation, Writing - review & editing. **Sandra L. McLellan:** Resources, Writing - review & editing, Supervision, Funding acquisition. **Melissa K. Schussman:** Data Curation, Methodology, Validation, Writing - review & editing. **Michelle Ammerman:** Data Curation, Methodology, Validation, Writing - review & editing. **Krista R. Wigginton:** Resources, Writing - review & editing, Supervision, Funding acquisition. **Kevin M. Bakker:** Data Curation, Writing - review & editing, Funding acquisition. **Alexandria B. Boehm:** Conceptualization, Methodology, Validation, Investigation, Formal analysis, Data curation, Writing - original draft, Supervision, Funding acquisition, Project Administration.

