## Supplemental Information for "SARS-CoV-2 RNA is enriched by orders of magnitude in solid relative to liquid wastewater at publicly owned treatment works"

### SARS-CoV-2 RNA concentrations in matched wastewater influent and settled solids at five publicly owned treatment works

**Laboratory participation**

Solid samples were processed by the labs at Stanford University (SB, JI, OC), University of Michigan (AA), and Verily (OS). Influent samples were processed by the lab at Southern California Coastal Water Research Project (SB, OC), University of Michigan (AA), UC Berkeley (OS), and University of Wisconsin-Milwaukee (JI).

**Laboratory specific procedures - additional details**

*Solids*

Frozen samples were thawed at 4°C for 12-36 hours and processed according to Wolfe et al.,[1] with modifications for samples from all POTW except OS which were processed exactly according to the publication.

40 mL of primary solids were centrifuged at 24,000xg for 30 minutes at 4°C and the supernatant was decanted. Approximately 0.5 g of the dewatered solids was dried at 110°C for up to 24 hours to determine its dry weight. For SB and OC, the dewatered solids were resuspended in DNA/RNA shield (Zymo Research, CA) in 15 mL falcon tubes to achieve 75 mg of solids (wet weight) per mL of the DNA/RNA shield, then stored in 4℃ for up to 48 hours until extraction. 5.25 µL of bovine coronavirus (BCoV) (Calf-guard Cattle Vaccine, PBS Animal Health, OH) was spiked into all samples a few hours before homogenization. For JI, the dewatered solids were stored at 4°C for up to 48 hours, and a mixture of DNA/RNA shield and BCoV (1.5 µL of BCoV/mL shield) was used to resuspend the solids to 75 mg of solids (wet weight) per mL of the BCoV-spiked solution a few hours before extraction. For AA dewatered solids were suspended in BCoV-spiked solution at a concentration of 37.5 mg/l. These concentrations of solids in solution optimized sensitivity while reducing RT-PCR inhibition.[1]

For AA, 0.5 g of 0.5 mm silica/zirconia beads (Biospec Products, OK) were added to each sample and homogenized by shaking with a Biospec Mini-Beadbeater-96 (Biospec Products, OK). For all other POTW, 5/32" Stainless Steel Grinding Balls (OPS Diagnostics, NJ) were added to each sample and homogenized by shaking with a Geno/Grinder 2010 (Spex SamplePrep, NJ). After the homogenization step, samples were briefly centrifuged and 300 µL of the supernatant was used for each replicate; for OS 300 µL of homogenized sludge was used. For SB, OC, and JI, RNA was extracted from duplicate aliquots per sample; for AA, RNA was extracted from triplicate aliquots per sample; for OS, RNA was extracted from ten replicate aliquots as described by Wolfe et al.[1] Extractions were done using the Chemagic 360 and the Chemagic™ Viral DNA/RNA 300 Kit H96 (Perkin Elmer, MA). Inhibitors were removed with Zymo OneStep-96 PCR Inhibitor Removal Kits (Zymo Research, CA) before storing the RNA in -80℃ for up to 78 days. Extraction negative controls (water) were extracted using the same protocol. Extraction positive controls, BCoV spiked in DNA/RNA shield, were extracted by adding 4 uL of Poly-A as carrier RNA.

Nucleic acids (NA) were quantified through one-step digital droplet (dd)RT-PCR for SARS-CoV-2 targets, BCoV, and Pepper Mild Mottle Virus (PMMoV). BioRad SARS-CoV-2 droplet digital

PCR kits were used with a BioRad QX200 AutoDG droplet digital PCR system (BioRad, CA). Full methods for OS solids are in Wolfe et al.[1]

For SB, OC, and JI, N1 and N2 were quantified using a duplex assay with undiluted NA template; each of the two replicate extractions were run in triplicate wells for a total of six wells per sample. Nine no-template controls (NTCs) were included on each plate. N1 and N2 positive controls were run in two wells per plate and consisted of NA from a nasopharynx swab of a high-titer patient from Stanford Hospital. For samples from the same POTWs, PMMoV and BCoV were quantified using a duplex assay with 1:100 diluted template; each of the two replicate extractions was run in one well for a total of two wells per sample. Four NTCs were included on each plate. Positive controls for BCoV (direct extraction of BCoV vaccine diluted to ~$10^6$ cp/mL) and PMMoV (synthetic DNA ultramer from IDT) were included in two wells each. Replicate wells were merged and processed in QuantaSoft and QuantaSoft Analysis Pro (BioRad, CA) to manually threshold and export data as described in Graham et al.[2]

For AA, N1 and N2 were quantified using a duplex assay with undiluted NA template; each of the three replicate extractions were run in one well for a total of three wells per sample. At least three wells of NTC were included on each plate, and N1 and N2 positive controls were run in three wells per plate (IDT plasmids). PMMoV and BCoV were quantified using a duplex assay with 1:100 diluted NA template; each of the three replicate extractions was run in one well for a total of three wells per sample. At least three wells of NTC were included on each plate; three wells of positive control (PMMoV synthetic DNA ultramer from IDT and BCoV spiked in water) were included on each plate, QuantaSoft and QuantaSoftAnalysis Pro (BioRad, CA) were used to manually threshold and export data.

The required number of droplets was 10,000 for individual wells; any samples with fewer droplets were rerun or not included in the final analysis. Merged wells with three or more positive droplets were deemed positive. For a plate to be included in further analysis, merged NTCs were required to have no more than two positive droplets. Any samples that did not return a value for PMMoV or BCoV were not included in the final analysis, assuming failed extraction. Six samples for JI were excluded based on these criteria.

*SB, OC influent*

Following the methods described in Steel et al.,[3] 500 mL of raw influent was acidified by adding 20% HCl to achieve pH of 3.5 or less. $MgCl_2$ was added to each sample bottle to a final concentration of 25 mM. Each sample was spiked with 150 µL of BCoV, including the filter blank (sterilized Phosphate Buffered Solution, Fisher BioReagents, MA). 20 mL of the samples were filtered through 0.45 µm pore size mixed cellulose ester HA filters (Millipore Sigma, MA) and stored at -80℃ for up to 2 months until NA extraction. For NA extraction, HA filters were added to Zymo BeadBashing beads to beat for a total of 2 minutes after spiking with armored Hep G (Asuragen, TX) as extraction control. After centrifuging, the supernatant was processed using BioMerieux Nucleic Extraction Kit (BioMerieux, NC) by following the protocols provided by the manufacturers. Extracted nucleic acid was stored at -80°C for up to 24 hours before analysis. Nucleic acids were quantified through one-step ddRT-PCR for SARS-CoV-2 (N1 and N2),

BCoV, and PMMoV. BioRad one-step RT-ddPCR Advanced Kit for Probes were used with a BioRad manual droplet generator and QX200 droplet digital PCR system (BioRad, CA). At least two technical replicates were quantified either undiluted or at a 1:2 dilution for SARS-CoV-2 targets and BCoV; PMMoV was quantified at a 1:10 dilution. However, if the concentration of the target gene was suspected to be low, four technical replicates were run. The positive controls were used one each plate for N1, N2 (IDT plasmids), BCoV and Hep G (1:1 mix of armored Hep G and 1:10 dilution of BCoV vaccine in water, heated to 75℃ for 3 minutes), and PMMoV (a previously extracted sewage sample). At least four no template controls (NTCs) were included on every plate. Technical replicates were merged and processed in QuantaSoft and QuantaSoft Analysis Pro using manual thresholding (BioRad, CA).

For a plate to be included in further analysis, merged NTCs were required to have no more than two positive droplets. The required number of droplets for merged wells was 10,000. Three or more positive droplets in a merged well after subtracting the number of positive droplets found in NTC were deemed positive. Each measurement had to have more than five negative droplets or was otherwise considered overloaded and was excluded from analysis or rerun. BCoV was used to calculate recovery throughout the entire process and Hep G was used as extraction and inhibition control. BCoV was used after 3 Jun 2020, so measurements before then do not have a BCoV recovery associated with the sample. Any samples with less than three droplets for Hep G, BCoV, or PMMoV were not included in the final analysis. BCoV recovery had to be above 3% or the sample was excluded or rerun.

*JI influent*

Feng et al.[4] provides the full methods used. The methods are similar to those used for OC and SB and are filtration based.

*AA influent*

PEG concentration method was used to extract nucleic acids, which were quantified using one-step ddRT-PCR for SARS-CoV-2 (N1 and N2), BCoV, and PMMoV.[5] BioRad SARS-CoV-2 droplet digital PCR kits were used with a BioRad QX200 AutoDG droplet digital PCR system (BioRad, CA). At least two technical replicates were quantified either undiluted for SARS-CoV-2 targets and BCoV; PMMoV was quantified at a 1:100 dilution. The positive controls were used three each plate for N1, N2, (gRNA from ATCC, ATCC VR-3276SD) and BCoV (BCoV spiked in water). At least three wells of NTC were included on each plate. The concentration per reaction was converted to copies per volume of wastewater using dimensional analysis.

*OS influent*

The sample collection, processing  and reverse transcription quantitative polymerase chain reaction (RT-qPCR) protocols were in development throughout the time period of sampling, as described by Kantor et al.[6] The major changes relevant to the collection of data are outlined throughout this section.

For each sample, 40 mL of raw wastewater was collected in a sterile centrifuge tube containing sodium chloride and buffer and shipped on ice to the lab at UC Berkeley. Samples were extracted and quantified within about three days of collection, as was previously determined to be adequate storage conditions and time for the extraction method.[7]

SARS-CoV-2 RNA was extracted directly from wastewater following the Sewage, Salt, Silica, and SARS-CoV-2 (4S) method,[7] with minor changes throughout the sampling period reflected in versions 2-4 of the protocol.[8] Extraction was completed without replication until 8 Dec 2020, after which extraction duplicates were processed for all samples. Bovilis® coronavirus (Merck Animal Health, NJ) was spiked into each sample and quantified as an extraction positive control in a subset of samples until 15 Mar 2021 when this procedure was extended to all samples.

Sample extracts underwent RT-qPCR targeting N1, PMMoV, BCoV, and VetMAX™ Xeno™ Internal Positive Control (Xeno). After March 15, 2021, a duplexed assay for PMMoV with BCoV replaced the individual assays. No-template controls, extraction negative controls, and standards on each plate were quantified in triplicate. Automatic thresholding on a Quant Studio 3 Real-Time qPCR system (ThermoFisher Scientific, MA) was used to determine Cq values (Design and analysis software v1.5.1) with thermocycling conditions in Table S9.

For the N1 assay, the limit of detection was assessed using the DNA standards to be 4 gene copies per reaction (gc/rxn). For BCoV and PMMoV, it was set at the bottom of the standard curve, which no samples fell below. Negative controls were all below the detection limit or had a higher Cq value than the lowest standard run. Individual standard curves were combined into master standard curves for each assay (Table S10) as described by Kantor et al.[6] RNA standards were used for N1 (Twist Bioscience, CA), PMMoV (IDT ultramer), and BCoV (IDT ultramer) assays until 11/4/20, after which DNA standards were used for N1 (2019-nCoV RUO kit), PMMoV (IDT gblock), and BCoV (IDT gblock). A detailed description of this process has been described previously by Kantor et al.[6] Outliers were assessed using a two-sided Grubbs test (alpha=0.05) on Cq triplicates.

RT-PCR inhibition was assessed by an internal positive control (Xeno) until further study found this method inadequate compared to serial dilution.[9] For samples collected after 11/13/20, serial dilution was completed to assess for inhibition by comparing 1x and 5x diluted samples.[2,9] The higher (adjusted) value between these dilutions was used in this study.

**Dimensional Analysis**

*Solids*

In order to convert from X copies/uL from ddPCR to Y copies/g dry weight, the following equation was used for all samples.

$$X\frac{copies}{\mu L\ rxn} \times \frac{Volume\ of\ rxn\ (\mu L)}{Volume\ of\ template\ in\ rxn\ (\mu L)} \times dilution\ factor \times \frac{Volume\ of\ eluent\ from\ extract\ (\mu L)}{Wet\ mass\ of\ solids\ in\ extract\ (g)} \times \%\ solids\ of\ sample\ = Y\frac{copies}{g\ dry\ weight}$$

*Influent*

For SB, OC, and JI, the following equation was used to convert copies/uL from ddPCR to copies/L wastewater.

$$X\frac{copies}{uL\ rxn} \times \frac{Volume\ of\ rxn\ (\mu L)}{Volume\ of\ template\ in\ rxn\ (\mu L)} \times dilution\ factor \times \frac{Volume\ of\ eluent\ from\ extract\ (\mu L)}{Volume\ of\ lysate\ for\ extraction\ (mL)} \times \frac{Volume\ of\ lysis\ buffer\ added\ (mL)}{Volume\ filtered\ (mL)} = Y\frac{copies}{mL\ wastewater}$$

For OS, the following equation was used for all samples with a dilution factor of 1 or 5.

$$X\frac{copies}{rxn} \times \frac{rxn}{Volume\ of\ template\ (\mu L)} \times \frac{Volume\ of\ eluent\ from\ extract\ (\mu L)}{Weight\ of\ sample\ (mg)} \times \frac{1\ mg}{1\ uL} \times \frac{1000\ uL}{1\ mL} \times dilution\ factor = Y\frac{copies}{mL\ wastewater}$$

For AA,

$$X\ \frac{copies}{rxn} \times \frac{rxn}{Volume\ of\ template\ (\mu L)} \times \mu L\ eluent\ from\ extract \times \frac{Volume\ of\ final\ concentrated\ volume\ (mL)}{Volume\ of\ concentrated\ sample\ used\ in\ extraction\ (mL)} \times \frac{1}{Volume\ of\ initial\ wastewater\ (mL)} = Y\frac{copies}{mL\ wastewater}$$

**Lower measurement limit.**

AA solids and OS solids lower measurement limit was calculated by the respective lab and reported based on the concentration that would be obtained with three positive droplets in ddPCR. For OS influent, lower measurement limit was assessed using DNA standards with qPCR, as described above. For all other data sets, the lower measurement limit for individual samples were calculated based on the three positive droplet cut-off and the average was reported. Additionally, for AA influent, different amounts of water was used when resuspending the viral PEG pellet, resulting in a unique effective volume for all samples. For JI influent, as the procedure evolved, the effective volume associated with the sample also differed. In both of these cases, an average of the lower measurement limit was calculated and reported. Lower measurement limit for data sets in this study are shown in Table S4.

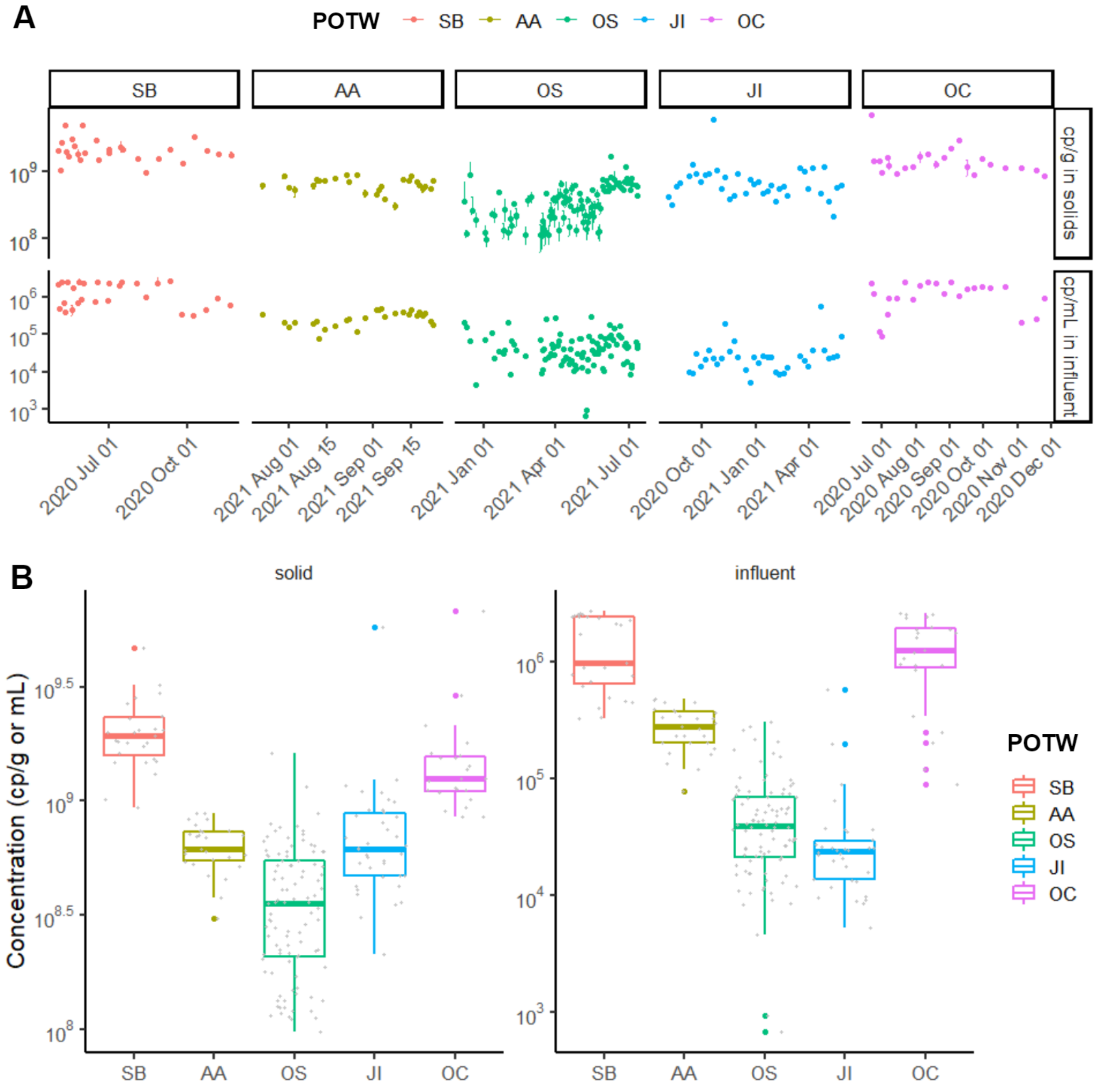


Figure S1. PMMoV concentration for each of the POTW sampled. **A.** Time series of PMMoV measured in solids (top) and influent (bottom). Standard deviations are plotted as error bars on all data points except OS and JI influent, as they were not reported. Some of the error bars are too small to be seen in the figure. **B.** Boxplot showing distribution of PMMoV concentration for each POTW. On the left is solid concentrations (cp/g dry weight) and on the right is influent (cp/mL wastewater). The line through the box represents the median, and the top and bottom of the box represent 75th and 25th percentile, respectively. The top and bottom whiskers show 1.5 times the upper and lower interquartile range, respectively. Data beyond this range is plotted in colored symbols. Individual data points are shown in grey.

**A. N1 or N**

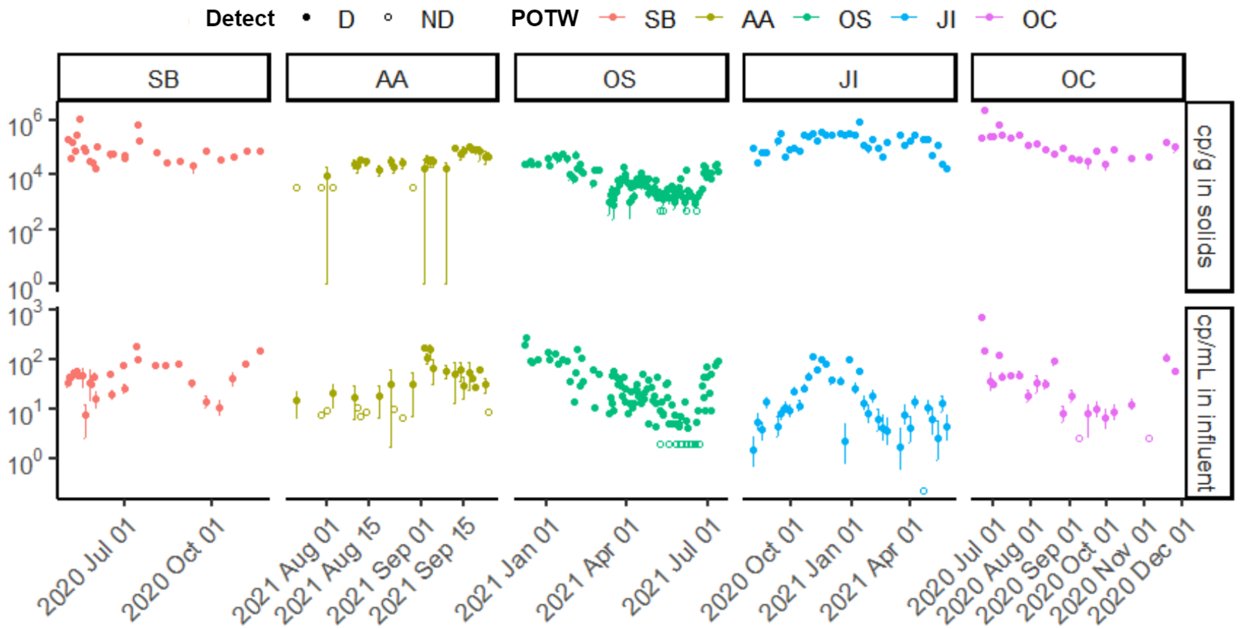


**B. N2**

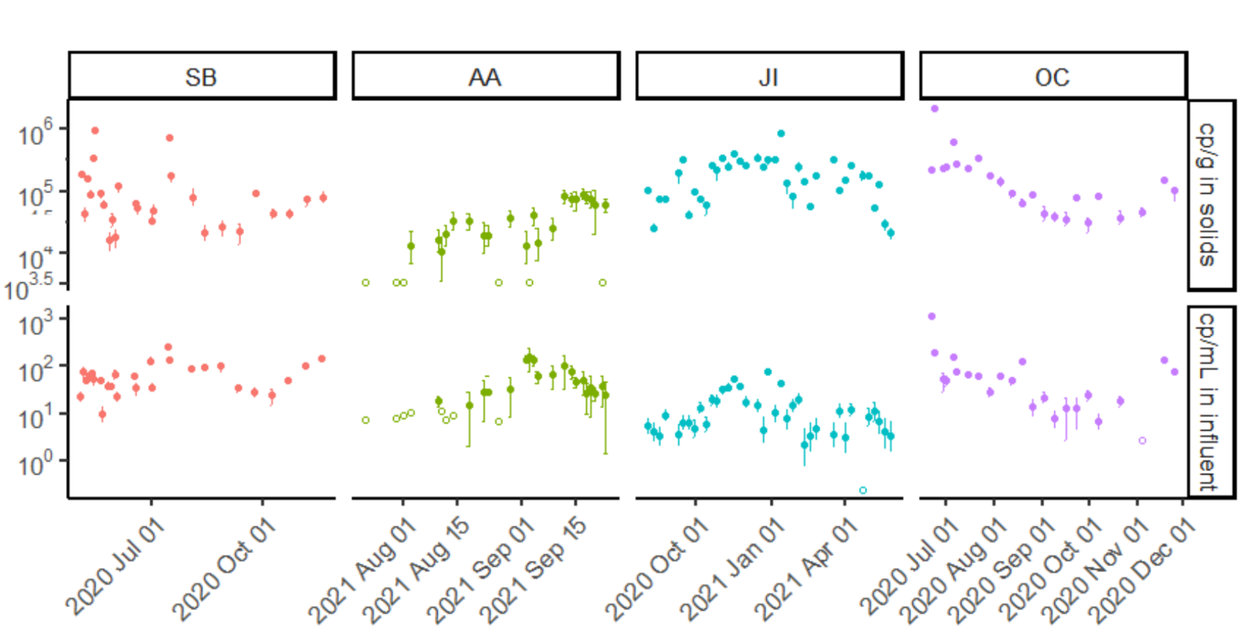


Figure S2. SARS-CoV-2 target concentrations for each POTW. **A.** Time series of N1 or N measured in solids (top) and influent (bottom). Standard deviations are plotted as error bars on all data points except OS influent, as they were not available for all samples. Samples above the lower measurement limit are shown as filled circles. Samples that resulted in ND, shown as empty circles, were substituted with a value half of the lower measurable limit. **B.** Time series of N2 measured in solid (top) and influent (bottom). Since N2 was not measured in OS, only four POTWs are shown. Standard deviations are plotted as error bars on all data points. Samples above the lower measurement limit are shown as filled circles. Samples that resulted in ND, shown as empty circles, were substituted with a value half of the lower measurable limit.

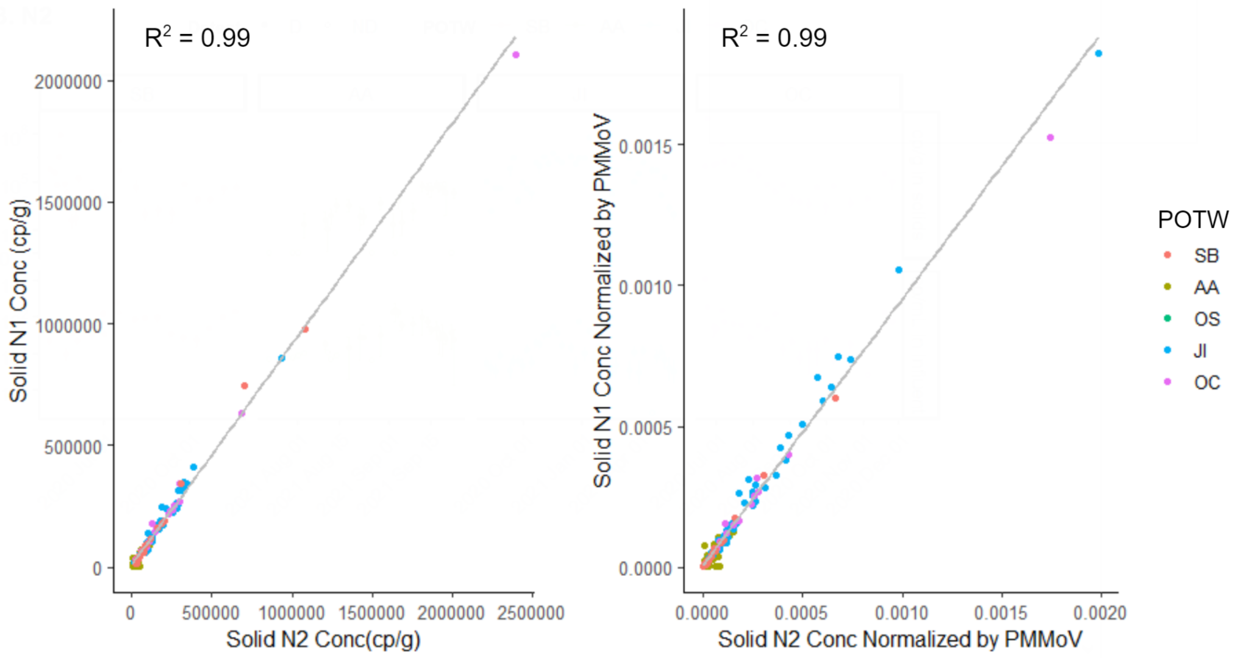


Figure S3. Pairwise linear regression between N1 and N2 solid concentrations from samples that detected both N1 and N2 (SB, AA, JI, OC). On the left are raw concentrations ($R^2$ = 0.99, slope = 1.1, p-value < $10^{-15}$), and on the right are concentrations normalized by PMMoV ($R^2$ = 0.99, slope = 1.0, p-value < $10^{-15}$).

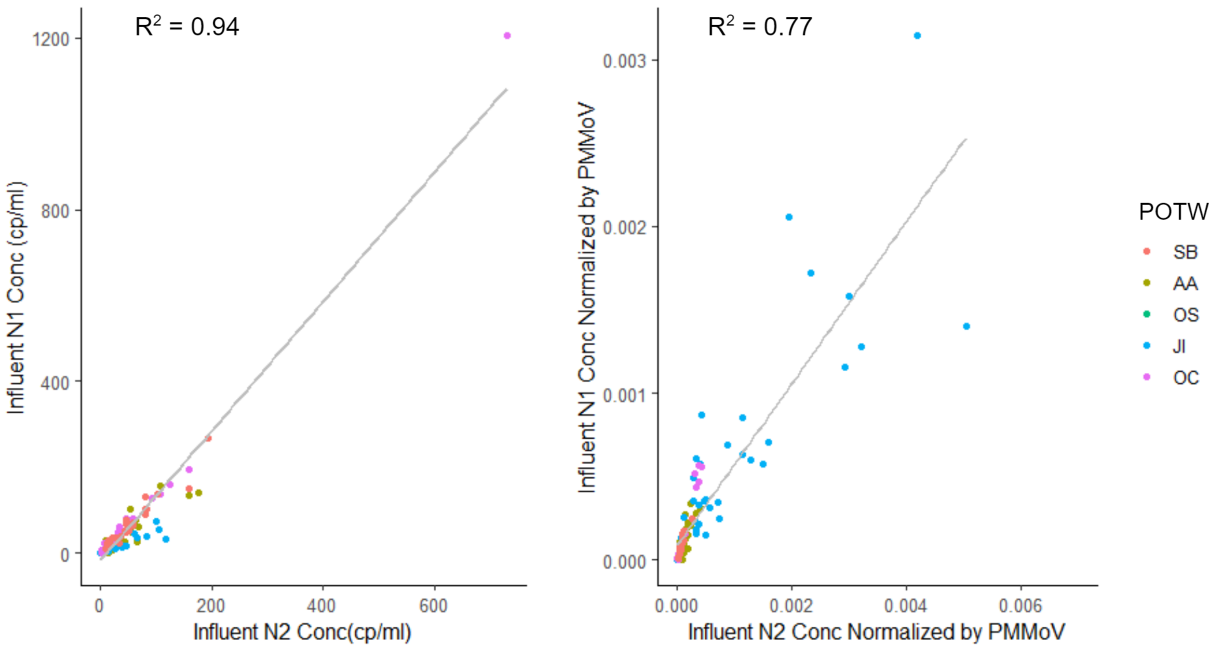


Figure S4. Pairwise linear regression between N1 and N2 influent concentrations from samples that detected both N1 and N2 (SB, AA, JI, OC). On the left are raw concentrations ($R^2$ = 0.93, slope = 0.6, p-value < $10^{-16}$) and on the right are concentrations normalized by PMMoV ($R^2$ = 0.77, slope = 1.6, p-value < $10^{-16}$).

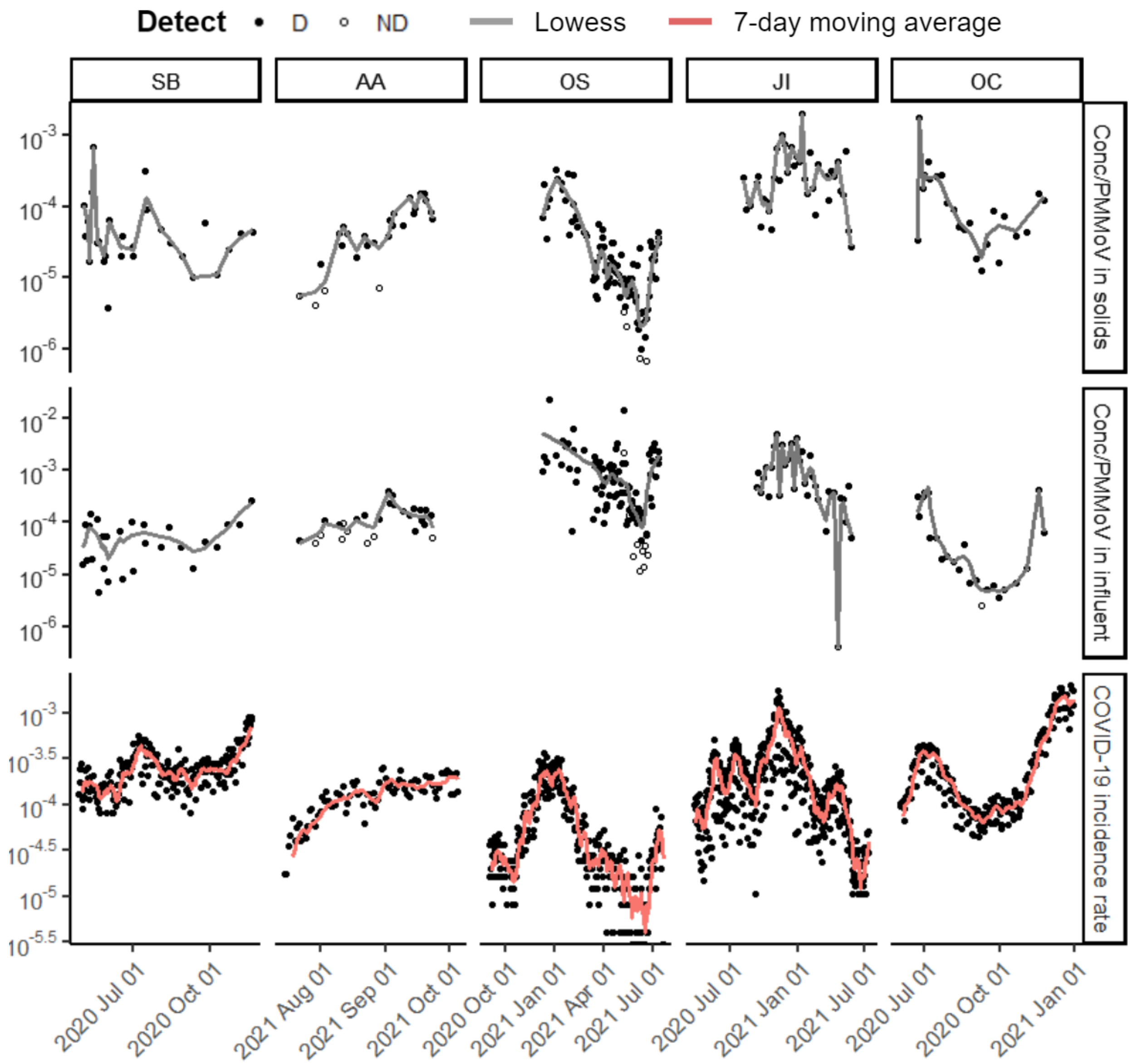


Figure S5. Time series of (top to bottom) SARS-CoV-2 targets N1 or N measured in solids (cp/g dry weight) normalized by PMMoV, concentration measured in influent (cp/mL) normalized by PMMoV and laboratory-confirmed SARS-CoV-2 incidence rate for each of the five POTWs over their respective duration of sample collection. N was measured for OS solids and N1 for all other data sets. Points represent individual data points. Samples above the lower measurement limit are shown as filled circles. Samples that resulted in ND, shown as empty circles, were substituted with a value half of the lower measurable limit. Lines for solid and influent are locally weighted scatterplot smoothing (lowess) with value of $\alpha$ that minimizes the residual for each dataset (Table S6).[9] Lines for COVID-19 incidence rates are 7-day centered smoothed averages.

Table S1. Sampling procedures associated with each plant for both influent and primary settled solids.

| | Influent | Primary Settled Solids |
|---|---|---|
| SB | Flow-weighted 24 hour composites | One grab sample taken in the morning |
| AA | Time-weighted 24 hour composites collected every 15 min | One grab sample, time of collection varied |
| OS | Time-weighted 24 hour composite collected every hour | One grab sample taken in the morning |
| JI | Flow-weighted 24 hour composites | Six grab samples collected every 4 hours composited |
| OC | Time-weighted 24 hour composites collected every 30 minutes | Two grab samples collected at 7am and 7pm composited |

Table S2. Additional information about each POTW. Chemical additions during the wastewater treatment process upstream of sampling points were noted here. The primary clarifier residence time was an estimate for residence time of the settled solids provided by the operators at each respective plant based on the hydraulic residence time. NA means not available, but solids residence times are usually less than 12 hours in primary clarifiers.

| POTW | Chemical Additions | Estimated primary clarifier residence time (hr) |
|---|---|---|
| SB | - No chemical additions | 3-6 |
| AA | - No chemical additions | NA |
| OS | - No chemical additions | 3-6 |
| JI | - Ferric chloride for odor control and improved settling efficiency on occasion | 1-2 |
| OC | - Hydrogen peroxide to influent for odor & corrosion control<br>- Ferric chloride and anionic polymer to primary clarifier for improved settling efficiency | 2-4 |

Table S3. Sample storage condition until preanalytical step. <1 day indicates that the sample was processed on the day of collection.

| POTW | Sample | Temperature | Days stored at specified temperature | |
|---|---|---|---|---|
| | | | Min | Max |
| AA | Influent | 4℃ | <1 | 3 |
| | Solids | 4℃ | <1 | 7 |
| SB | Influent | 4℃ | <1 | 3 |
| | Solids | -80℃ | 173 | 371 |
| OS | Influent | 4℃ | <1 | 3 |
| | Solids | 4℃ | <1 | <1 |
| JI | Influent | 4℃ | 1 | 1 |
| | Solids | -80℃ | 28 | 428 |
| OC | Influent | 4℃ | <1 | 3 |
| | Solid | -80℃ | 159 | 329 |

Table S4. Estimated lower measurement limit  for both solids and influent samples in each POTW. For influent, where samples had variable volumes processed, and therefore different lower measurement limits, the average of sample-specific lower measurement limits are reported.

| | SB | AA | OS | JI | OC |
|---|---|---|---|---|---|
| Solid (cp/g) | 4300 | 6,800 | 900 | 3000 | 3000 |
| Influent (cp/ml) | 27 | 19 | 4 | 0.44 | 5.2 |

Table S5. Ratio of PMMoV concentrations in matched solid to influent samples in each POTW listed as a row. Number of matched samples and minimum, median, and maximum ratios calculated for the plants are reported. Note that some samples did not have PMMoV measured so the number of samples (N) in this table is different for PMMoV and SARS-CoV-2 N1 or N targets.

| Plant | N | Min | Median | Max |
|---|---|---|---|---|
| SB | 27 | $6x10^2$ | $1x10^3$ | $1x10^4$ |
| AA | 27 | $8x10^2$ | $2x10^3$ | $9x10^3$ |
| OS | 96 | $6x10^2$ | $9x10^3$ | $3x10^5$ |
| JI | 34 | $8x10^2$ | $3x10^4$ | $3x10^5$ |
| OC | 23 | $4x10^2$ | $1x10^3$ | $1x10^4$ |

Table_S6. $\alpha$ values used to plot lowess lines in Figure 2 (in column N1/PMMoV or N/PMMoV) and Figure S5 (in column N1 or N). Plants and matrices are listed as rows. These $\alpha$ values were chosen to minimize the residual for each dataset.

| Plant | Matrix | N1 or N | N1/PMMoV or N/PMMoV |
|---|---|---|---|
| SB | Solids | 0.221 | 0.221 |
| | Influent | 0.221 | 0.221 |
| AA | Solids | 0.174 | 0.174 |
| | Influent | 0.221 | 0.221 |
| OS | Solids | 0.174 | 0.112 |
| | Influent | 0.314 | 0.205 |
| JI | Solids | 0.128 | 0.128 |
| | Influent | 0.128 | 0.143 |
| OC | Solids | 0.252 | 0.252 |
| | Influent | 0.205 | 0.252 |

Table S7. Median Kendall's tau correlation between matched solid and influent SARS-CoV-2 concentration. 1000 instances of Kendall's tau were calculated by bootstrapping upper and lower bounds for measured concentration of SARS-CoV-2 RNA. Confidence intervals were not available for all OS influent measurements, and therefore Kendall's tau was calculated with raw data points. Kendall's tau was calculated with N1 or N wastewater concentration and with values normalized by PMMoV. Empirical p-value was below 0.005 unless otherwise stated in parentheses.

| Plant | N1 or N | N1/PMMoV or N/PMMoV |
|---|---|---|
| All | 0.22 | 0.11 |
| SB | 0.12 | 0.01 (p-value = 0.28) |
| AA | 0.20 | 0.22 |
| OS | 0.46 | 0.41 |
| JI | 0.33 | 0.27 |
| OC | 0.54 | 0.52 |

Table S8. Linear regression coefficients between $log_{10}$-transformed N1 or N concentrations of matched influent and solids: Y = mx+b where y = $log_{10}$-transformed solids concentrations, m = slope, b = intercept and x = $log_{10}$-transformed influent concentration. The error on m and b represents standard error for the calculated coefficients. $R^2$ and p-value are provided for completeness (Kendall's tau is used to assess association, see Table S7).

| POTW | m | b | $R^2$ | p-value |
|---|---|---|---|---|
| All | 0.47 ± 0.10 | 3.71 ± 0.14 | 0.10 | $< 10^{-5}$ |
| SB | 0.32 ± 0.26 | 4.34 ± 0.43 | 0.02 | 0.2174 |
| AA | 0.35 ± 0.21 | 3.90 ± 0.31 | 0.07 | 0.10 |
| OS | 0.63 ± 0.07 | 2.89 ± 0.10 | 0.41 | $<10^{-12}$ |
| JI | 0.26 ± 0.10 | 4.89 ± 0.12 | 0.13 | 0.01 |
| OC | 0.55 ± 0.13 | 4.29 ± 0.20 | 0.45 | $<10^{-3}$ |

Table S9. PCR cycling conditions used for target quantification. The matrix and POTW they were used for is provided in the first column. For dd RT-PCR methods, after cycling was complete, plates were either analyzed immediately or placed in 4℃ until analysis with the plate reader. The superscript in the sample description column provides a reference for the assay conditions.

| **Sample description** | **Assay** | **Step** | **Cycle #s** | **Temp (℃)** | **Time (min)** |
|---|---|---|---|---|---|
| Solids JI, SB, OC and AA; and influent AA | SARS-CoV-2_N1/N2 | Reverse transcription | 1 | 50 | 60 |
| | | Enzyme activation | 1 | 95 | 10 |
| | | Denaturing | 40 | 94 | 0.5 |
| | | Annealing | | 55 | 0.5 |
| | | Enzyme deactivation | 1 | 98 | 10 |
| | | Droplet stabilization | 1 | 4 | 30 |
| | PMMoV/BCoV | Reverse transcription | 1 | 50 | 60 |
| | | Enzyme activation | 1 | 95 | 10 |
| | | Denaturing | 40 | 94 | 0.5 |
| | | Annealing | | 56 | 0.5 |
| | | Enzyme deactivation | 1 | 98 | 10 |
| | | Droplet stabilization | 1 | 4 | 30 |
| Solids OS[1] | SARS-CoV-2 assay | Reverse transcription | 1 | 50 | 60 |
| | | Enzyme activation | 1 | 95 | 5 |
| | | Denaturing | 40 | 95 | 0.5 |
| | | Annealing | | 59 | 0.5 |

| | | | | | |
|---|---|---|---|---|---|
| | | Enzyme deactivation | 1 | 98 | 10 |
| | | Indefinite hold | 1 | 4 | ∞ |
| | PMMoV/BCoV | Reverse transcription | 1 | 50 | 60 |
| | | Enzyme activation | 1 | 95 | 5 |
| | | Denaturing | 40 | 95 | 0.5 |
| | | Annealing | | 56 | 0.5 |
| | | Enzyme deactivation | 1 | 98 | 10 |
| | | Indefinite hold | 1 | 4 | ∞ |
| Influent SB and OC | All | Reverse transcription | 1 | 50 | 60 |
| | | Enzyme activation | 1 | 95 | 10 |
| | | Denaturing | 40 | 95 | 0.5 |
| | | Annealing | | 58 | 0.5 |
| | | Enzyme deactivation | 1 | 98 | 10 |
| | | Hold | 1 | 12 | 20 |
| Influent JI[4] | SARS-CoV-2_N1/N2 & BCoV | Reverse transcription | 1 | 50 | 60 |
| | | Enzyme activation | 1 | 95 | 10 |
| | | Denaturing | 40 | 94 | 0.5 |
| | | Annealing | | 55 | 1 |
| | | Enzyme deactivation | 1 | 98 | 10 |
| | | Hold | 1 | 4 | 30 |

| | | | | | |
|---|---|---|---|---|---|
| | PMMoV | Reverse transcription | 1 | 50 | 60 |
| | | Enzyme activation | 1 | 95 | 10 |
| | | Denaturing | 40 | 94 | 0.5 |
| | | Annealing | | 60 | 1 |
| | | Enzyme deactivation | 1 | 98 | 10 |
| | | Hold | 1 | 4 | 30 |
| Influent OS[7] | All | Uracil-DNA glycosylase incubation | 1 | 25 | 2 |
| | | Reverse transcription | 1 | 50 | 15 |
| | | Enzyme activation | 1 | 95 | 2 |
| | | Denaturing | 45 | 95 | 0.05 |
| | | Annealing | | 55 | 0.5 |

Table S10: Master standard curves used to calculate quantities for each assay as applied in the 4S method for OS influent. The Target is the assay target. The standard curve minimum and maximum are the lowest and highest concentration standard used in generating the standard curve, respectively. Slope is the slope, m, of the standard curve; b is the y-intercept of the standard curve where y = mx + b and y is $\log_{10}$ gc/rxn and x is Cq.

| **Target** | **Standard curve minimum (cp/rxn)** | **Standard curve maximum (cp/rxn)** | **slope** | **intercept** | **$R^2$** |
|---|---|---|---|---|---|
| N1 | 5 | $10^5$ | -3.37 | 38.5 | 0.92 |
| PMMoV | $10^2$ | $10^8$ | -3.31 | 40.5 | 0.88 |

Table S11. Primer and probe sequences used in the study. The matrix and POTW they were used for is provided in the first column along with a reference. The superscript in the sample description column provides a reference for the sequences.

| Sample description | Amplicon | Forward Primer | Reverse Primer | Probe Sequence |
|---|---|---|---|---|
| Solids JI, SB, OC and AA; and influent AA | SARS-CoV-2_N1 | GACCCCAAA ATCAGCGAAAT | TCTGGTTACTGC CAGTTGAATCTG | FAM-ACCCCGCATTAC GTTTGGTGGACC -IBFQ |
| | SARS-CoV-2_N2 | TTACAAACATTG GCCGCAAA | GCGCGACATTC CGAAGAA | HEX-ACAATTTGCCCC CAGCGCTTCAG-IBFQ |
| | BCoV | CTGGAAGTTGGT GGAGTT | ATTATCGGCCTA ACATACATC | CCTTCATATCTA TACACATCAAGT TGTT |
| | PMMoV | GAGTGGTTTGAC CTTAACGTTTGA | TTGTCGGTTGCA ATGCAAGT | FAM-CCTACCG AAGCAAATG-MGB-NFQ |
| Solids OS[1] | SARS-CoV-2_N | CATTACGTTTGG TGGACCCT | CCTTGCCATGTT GAGTGAGA | FAM/ZEN-CGCGATCAAAAC AACGTCGG-IBFQ |
| | BCoV | CTGGAAGTTGGT GGAGTT | ATTATCGGCCTA ACATACATC | ATTATCGGCCTA ACATACATC |
| | PMMoV | GAGTGGTTTGAC CTTAACGTTTGA | GAGTGGTTTGAC CTTAACGTTTGA | HEX/ZEN-CCTACCGAAGCA AATG-IBFQ |
| Influent SB and OC | SARS-CoV-2_N1 | GACCCCAAAATC AGCGAAAT | TCTGGTTACTGC CAGTTGAATCTG | FAM-ACCCCGCATTAC GTTTGGTGGACC -BHQ1 |
| | SARS-CoV-2_N2 | TTACAAACATTG GCCGCAAA | GCGCGACATTC CGAAGAA | SUN-ACAATTTGCCCC CAGCGCTTCAG-BHQ1 |
| | Hep-G | GGCCAAAAGGT GGTG | GACGAGCCTGA CGTCG | FAM-TCCCTCTGG-ZEN-CGCTTGTGGC-3IABkFQ |

| | | | | |
|---|---|---|---|---|
| | BCoV | C+TGGAAGTTGG TGGAGTT | ATTATCGG+CCT AACATAC+ATC | HEX- ACCCAGAAA- ZEN- CAAACAACTTGA TGTGTATAGATA TGAA-3IABkFQ |
| | PMMoV | GAGTGGTTTGAC CTTAACGTTGA | TTGTCGGTTGCA ATGCAAGT | HEX- CCTACCGAAGCA AATG-3IABkFQ |
| Influent JI[4] | SARS-CoV-2_N1 | GACCCCAAAATC AGCGAAAT | TCTGGTTACTGC CAGTTGAATCTG | FAM- ACCCCGCAT- ZEN- TACGTTTGGTGG ACC-IABkFQ |
| | SARS-CoV-2_N2 | TTACAAACATTG GCCGCAAA | GCGCGACATTC CGAAGAA | HEX- ACAATTTGCCCC CAGCGCTTCAG- BHQ1 and HEX- ACAATTTGC- ZEN- CCCCAGCGCTT CAG-IABkFQ |
| | BCoV | CTGGAAGTTGGT GGAGTT | ATTATCGGCCTA ACATACATC | FAM- CCTTCATAT- ZEN- CTATACACATCA AGTTGTT-IA BkFQ |
| | PMMoV | GAGTGGTTTGAC CTTAACGTTGA | TTGTCGGTTGCA ATGCAAGT | FAM- CCTACCGAAGCA AATG-MGBNFQ |
| Influent OS[7] | SARS-CoV-2_N1 | GACCCCAAAATC AGCGAAAT | TCTGGTTACTGC CAGTTGAATCTG | FAM- ACCCCGCATTAC GTTTGGTGGACC - ZEN/IBFQ |
| | BCoV | CTGGAAGTTGGT GGAGTT | ATTATCGGCCTA ACATACATC | FAM- CCTTCATATCTA TACACATCAAGT TGTT- ZEN/IBFQ |

| | | | | |
|---|---|---|---|---|
| | PMMoV | GAGTGGTTTGACCTTAACGTTTGA | TTGTCGGTTGCAATGCAAGT | FAM-CCTACCGAAGCAAATG-ZEN/IBFQ |

#### References


1 M. K. Wolfe, A. Topol, A. Knudson, A. Simpson, B. White, D. J. Vugia, A. T. Yu, L. Li, M. Balliet, P. Stoddard, G. S. Han, K. R. Wigginton and A. B. Boehm, High-Frequency, High-Throughput Quantification of SARS-CoV-2 RNA in Wastewater Settled Solids at Eight Publicly Owned Treatment Works in Northern California Shows Strong Association with COVID-19 Incidence, *mSystems*, , DOI:10.1128/mSystems.00829-21.

2 K. E. Graham, S. K. Loeb, M. K. Wolfe, D. Catoe, N. Sinnott-Armstrong, S. Kim, K. M. Yamahara, L. M. Sassoubre, L. M. Mendoza Grijalva, L. Roldan-Hernandez, K. Langenfeld, K. R. Wigginton and A. B. Boehm, SARS-CoV-2 RNA in Wastewater Settled Solids Is Associated with COVID-19 Cases in a Large Urban Sewershed, *Environ. Sci. Technol.*, 2021, **55**, 488–498.

3 J. A. Steele, A. G. Zimmer-Faust, J. F. Griffith and S. B. Weisberg, *Sources of variability in methods for processing, storing, and concentrating SARS-CoV-2 in influent from urban wastewater treatment plants*, Public and Global Health, 2021.

4 S. Feng, A. Roguet, J. S. McClary-Gutierrez, R. J. Newton, N. Kloczko, J. G. Meiman and S. L. McLellan, Evaluation of Sampling, Analysis, and Normalization Methods for SARS-CoV-2 Concentrations in Wastewater to Assess COVID-19 Burdens in Wisconsin Communities, *ACS EST Water*, 2021, **1**, 1955–1965.

5 M. T. Flood, N. D'Souza, J. B. Rose and T. G. Aw, Methods Evaluation for Rapid Concentration and Quantification of SARS-CoV-2 in Raw Wastewater Using Droplet Digital and Quantitative RT-PCR, *Food Environ. Virol.*, 2021, **13**, 303–315.

6 R. S. Kantor, H. D. Greenwald, L. C. Kennedy, A. Hinkle, S. Harris-Lovett, M. Metzger, M. M. Thornton, J. M. Paluba and K. L. Nelson, *Operationalizing a routine wastewater monitoring laboratory for SARS-CoV-2*, Epidemiology, 2021.

7 O. N. Whitney, L. C. Kennedy, V. B. Fan, A. Hinkle, R. Kantor, H. Greenwald, A. Crits-Christoph, B. Al-Shayeb, M. Chaplin, A. C. Maurer, R. Tjian and K. L. Nelson, Sewage, Salt, Silica, and SARS-CoV-2 (4S): An Economical Kit-Free Method for Direct Capture of SARS-CoV-2 RNA from Wastewater, *Environ. Sci. Technol.*, 2021, **55**, 4880–4888.

8 O. N. Whitney, B. Al-Shayeb, A. Crits-Cristoph, M. Chaplin, V. Fan, H. Greenwald, A. Hinkle, R. Kantor, L. Kennedy, A. Maurer, R. Tjian, K. L. and U. Berkeley, 2020.

9 H. D. Greenwald, L. C. Kennedy, A. Hinkle, O. N. Whitney, V. B. Fan, A. Crits-Christoph, S. Harris-Lovett, A. I. Flamholz, B. Al-Shayeb, L. D. Liao, M. Beyers, D. Brown, A. R. Chakrabarti, J. Dow, D. Frost, M. Koekemoer, C. Lynch, P. Sarkar, E. White, R. Kantor and K. L. Nelson, Tools for interpretation of wastewater SARS-CoV-2 temporal and spatial trends demonstrated with data collected in the San Francisco Bay Area, *Water Res. X*, 2021, **12**, 100111.
